# Climatic drivers of seasonal dynamics for Respiratory Syncytial Virus (RSV) in Antananarivo, Madagascar, 2011-2021

**DOI:** 10.1101/2024.02.03.24302203

**Authors:** Tsiry Hasina Randriambolamanantsoa, Norosoa Harline Razanajatovo, Hafaliana Christian Ranaivoson, Laurence Randrianasolo, Joelinotahiana Hasina Rabarison, Helisoa Razafinmanjato, Arvé Ratsimbazafy, Danielle Aurore Doll Rakoto, Jean-Michel Heraud, Vincent Lacoste, Cara E. Brook

## Abstract

**Introduction:** Respiratory Syncytial Virus (RSV) is a primary source of acute lower respiratory tract infection (ALRTI), the leading cause of death in children under five. Over 99% of RSV-attributed deaths occur in low-income countries, including Madagascar. RSV transmission is linked to climate, driving highly seasonal dynamics.

**Methods:** We used generalized additive models (GAMs) to identify correlates of reported RSV infections in Antananarivo, Madagascar from January 2011-December 2021, then fit catalytic models to cumulative age-structured incidence to estimate age-specific force of infection (FOI). We fit a time series Susceptible-Infected-Recovered (TSIR) model to the dataset to estimate weekly RSV transmission, then evaluated associations with precipitation, humidity, and temperature using generalized linear models. We used GAMs to quantify interannual trends in climate and assess whether significant deviations in RSV burden occurred in years representing climatic anomalies.

**Results:** Reported RSV infections in Antananarivo were significantly associated with patient ages ≤2 years. Highest FOI was estimated in patients ≤1 year, with transmission declining to near-zero by age five before rising in older (60+) cohorts. TSIR models estimated a January–February peak in RSV transmission, which was strongly positively associated with precipitation and more weakly with temperature but negatively related to relative humidity. Precipitation, humidity, and temperature all increased across the study period in Antananarivo, while reported RSV infections remained stable. Significant deviations in RSV burden were not associated with clear climate anomalies.

**Conclusions:** Stable rates of reported RSV infections in Antananarivo across the past decade may reflect contrasting impacts of elevated precipitation and increased humidity on transmission. If future climate changes yield more rapidly accelerating precipitation than humidity, this could accelerate RSV burden. Introduction of recently-developed public health interventions to combat RSV in low-income settings like Madagascar is essential to mitigating burden of disease (RSV), in particular any future climate-driven increases in transmission or severity.

**Key Messages:**

- *What is already known on this topic*: RSV is an important driver of acute lower respiratory tract infections, which represent the leading cause of mortality in children under five across the globe. RSV demonstrates highly seasonal dynamics, as its transmission is linked to climate.
- *What this study adds:* We quantified correlates of RSV infection and estimated the seasonal transmission rate for RSV from reported patient data in Antananarivo, Madagascar. We found that RSV transmission is primarily concentrated in very young children (≤1 year) in Antananarivo and positively associated with high precipitation and low humidity, which focus most transmission in Madagascar’s January-February rainy season.
- *How this study might affect research, practice, or policy:* Our study suggests that RSV burden may intensify with future climate change, particularly higher rainfall. We emphasize the high public health importance of accelerating the introduction of recently-developed mAbs (Monoclonal Antibody) and vaccination interventions to combat RSV to low-income settings like Madagascar.

## INTRODUCTION

Respiratory syncytial virus (RSV) is a highly contagious virus that primarily infects infants and young children. Worldwide, most children contract RSV before reaching two years of age (Andeweg *et al*. 2021). The virus infects the upper and lower respiratory tracts causing infections that range from common cold to bronchiolitis and pneumonia. Globally, acute lower respiratory tract infections (ALRTIs) are the leading cause of death in children under five (CU5) (Data 2019), and approximately one-fourth of ALRTIs are estimated to be attributable to RSV each year (Nair *et al*. 2010, Shi *et al*. 2017). Over 99% of CU5 fatalities from RSV-related ALRTIs occur in low-income countries (Nair *et al*. 2010, Shi *et al*. 2017). RSV can also affect older age groups and those immuno-compromised or with comorbidities such as asthma and other lung diseases, diabetes, and heart failure (Falsey *et al*. 2005, Haber 2018). In Madagascar, RSV remains the most common cause of ALRTI and a major cause of hospital admission in CU5 *(Razanajatovo et al. 2018)*. It is estimated that approximately 11 299 hospitalizations per year can be attributed to RSV in CU5 in Madagascar (Rabarison *et al*. 2019). Three years (2010-2013) of laboratory surveillance from Institut Pasteur de Madagascar (IPM) aimed at identifying the etiology of severe acute respiratory infections (SARI) showed that nearly 37.7% of samples were positive for RSV, with the vast majority of patients (81%) representing CU5 (Razanajatovo *et al*. 2018). A further 38.9% of SARI surveillance samples collected between 2018-2022 in Madagascar additionally tested positive for RSV, again with the majority concentrated in CU5 (Razanajatovo *et al*. 2022).

Given the globally significant impact of RSV on childhood morbidity and mortality, RSV has been the subject of numerous research studies across many countries (Shi *et al*. 2017, O’Brien *et al*. 2019, Li *et al*. 2022). Particular areas of focus have included standard reporting of disease burden, clinical features, and at-risk groups for severe infections—typically aimed at defining best practices for the monitoring of RSV infection in CU5 and avoiding the misuse of antibiotics (Linssen *et al*. 2021, Obolski *et al*. 2021, Li *et al*. 2022, Pinquier *et al*. 2023). In addition, considerable work has been devoted to quantification of seasonal transmission dynamics for RSV—in order to better predict the timing and magnitude of severe epidemics (Hogan *et al*. 2016, Baker *et al*. 2019, Wambua *et al*. 2022).

The seasonality of RSV circulation shows different patterns depending on geographic location, though most localities are characterized by a clear annual peak in transmission. In temperate countries, peak cases generally predominate during the colder months—from September to January in the Northern Hemisphere and from March to June in the Southern Hemisphere (Obando-Pacheco *et al*. 2018). In tropical and subtropical countries, the timing of the peak RSV season is more variable across locations, with some studies reporting highest RSV activity during peak rains (Sapin *et al*. 2001, Mathisen *et al*. 2009, Matthew *et al*. 2009) and others reporting elevated caseloads during warmest months of the year (Chew *et al*. 1998). Madagascar has been previously documented as an anomaly in the study of seasonal circulation for respiratory viruses, particularly influenza, for which transmission is highly irregular (Heraud *et al*. 2019). RSV circulation, however, is well defined, with burden concentrated in the first half of each calendar year and the peak in cases between February and March (Razanajatovo *et al*. 2018, Razanajatovo *et al*. 2022).

Climate is known to play an important role in driving RSV circulation across diverse geographic localities (Shi *et al*. 2017, Baker *et al*. 2019, Heraud *et al*. 2019) but has remained largely unexplored in Madagascar. Quantitative understanding of the climatic factors that drive intra-annual seasonality in RSV transmission is essential to predicting how RSV dynamics will respond to future interannual climatic changes. Here, we aimed to (1) identify statistical correlates of RSV infection from reported cases of respiratory infection in Antananarivo, Madagascar over the past decade, (2) estimate variation in the age-structured force of infection (FOI) for RSV cases in Antananarivo, (3) quantify the seasonality of RSV transmission across the study period, and (4) identify climatic variables associated with peak RSV transmission.

## METHODS

### Study location and setting

The influenza sentinel surveillance network (ISSN) in Madagascar has been operational since 1978 (Rasolofonirina 2003, Randrianasolo *et al*. 2010). The aim is to monitor the circulation of influenza viruses as well as other respiratory viruses of public health concern, such as RSV and, more recently, SARS-CoV-2. The ISSN is comprised of an ILI (Influenza-Like Illness) surveillance program that now involves 21 referral primary health care centers (CSB_R) and a SARI (Severe Acute Respiratory Infection) surveillance program in five selected hospitals. The ISSN is complemented via the monitoring of death certificate and mortality data collected from six districts in Antananarivo through the Bureau Municipal d’Hygiène recently renamed DEAH (Direction de l’Eau, l’Assainissement et l’Hygiène) (Rabarison *et al*. 2023)

For the present study, we used data collected from sentinel sites in Antananarivo, the capital city of Madagascar, over 11 years (from January 2011 – December 2021). Antananarivo is home to around three million inhabitants (Desa 2018) and hosts a tropical climate profile that is hot and rainy in summer (November – April) and cold and dry in winter (May – October). Sites used in our analysis included ILI three sites BHK: Ostie Behoririka (−18.90, 47.53), CSMI TSL: Centre de Santé Maternelle et Infantile de Tsaralalana (−18.91, 47.53), MJR: Dispensaire Manjakaray (−18.89, 47.53); and two SARI sites (CENHOSOA: Centre Hospitalier de Soavinandriana (−18.90, 47.545) and CHUMET (Centre Hospitalier Universitaire Mère-Enfant Tsaralalàna) (−18.91, 47.52). Respectively, each site serves a catchment population of: 28574 people (BHK); 43222 people (CSMI TSL), 8000 people (MJR), 89000 people (CENHOSOA), and 43222 people (CHUMET).

### Study subjects

All patients presenting to focal sites with ILI or SARI symptoms were enrolled. Details on case definitions and enrollment procedures have been previously published. Briefly, ILI classification criteria refer to patients of any age reporting with (a) a recorded temperature ≥38°C or a history of fever and (b) a cough of ≤10 days duration, who (c) do not require hospitalization. SARI patients report with these same symptoms but additionally require hospitalization (WHO 2013). For each enrolled patient, demographic, socio-economic, clinical, and epidemiological data were recorded in case report forms.

### Virus detection

Nasopharyngeal specimens were collected for each enrolled patient. After collection, samples were shipped daily to the Virology Unit at IPM where they were immediately processed or stored at 4°C until testing (a maximum of 2 days after collection, following WHO guidelines) (WHO 2002, WHO 2011). All procedures for the processing of biological samples have been previously described (Razanajatovo *et al*. 2011). Briefly, nasopharyngeal swabs were screened for influenza A and B, RSV, and, since March 2020, SARS-CoV-2, using discrete real-time RT-PCR assays (Supplementary Text S1). Only the SARI sentinel site of CENHOSOA supplied samples consistently across the entire study period, while most sites supplied samples intermittently for shorter durations throughout the 2011-2021 time series (Figure S1).

### Climate data

Meteorological data reporting total precipitation (mm), mean relative humidity (%), and mean temperature (^0^C) at daily intervals in Antananarivo, Madagascar across the study period were downloaded from the National Aeronautics and Space Administration (NASA) database (NASA) and summarized by week (summed for precipitation and averaged for humidity and temperature).

### Data analysis

#### RSV infection correlates from generalized additive models (GAMs)

We first aimed to identify correlates of RSV infection in patients reporting to study sites with ILI and SARI symptoms across the study period. To this end, we fit a series of Generalized Additive Models (GAMs) in the binomial family, incorporating a response variable of RSV infection outcome (0/1, indicating negative or positive) for all ILI and SARI patients tested in dataset across all reporting sites (Wood 2001). In the first GAM, we modeled all predictor variables as smoothing splines, with ‘day of year’ formatted as a cyclic cubic spline (to control for intra-annual seasonality in our dataset), ‘age’ formatted as a thinplate smoothing spline, and ‘sex’, ‘year’, and ‘reporting hospital’ formatted as factorial random effects (Table S1). We additionally fit a second GAM with ‘year’ formatted as a numeric thinplate smoothing spline to evaluate interannual patterns across the time series (Table S1). Finally, because age and sex data were only available for a subset (N=3242) of tested cases, we fit a third GAM in the binomial family to the full dataset of RSV test data (N=3432), incorporating only predictor variables of ‘day of year’ (as a cubic smoothing spline) and ‘year’ (as a random effect) (Table S1).

#### Estimation of age-structured force of infection (FOI) for reported RSV cases in Antananarivo

Building from age correlates of infection identified in GAMs, we next sought to quantify age-structured variation in the FOI for reported RSV cases across the study period. Methods for estimating FOI from age-stratified serological data for single-strain immunizing pathogens are well-established (Muench 1959, Heisey *et al*. 2006, Long *et al*. 2010, Pomeroy *et al*. 2015), and prior work has successfully adapted them for application to age-structured incidence data, in lieu of serology (Grenfell *et al*. 1985). Assuming individuals are born susceptible and immunity following infection is lifelong, the standard ‘catalytic’ model describes the cumulative probability (*P*(*a*)) of encountering infection by age *a* as:

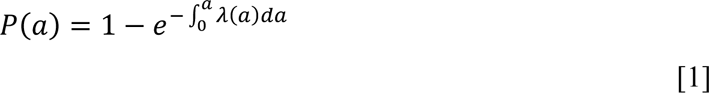

where *λ*(*a*) is the age-specific force of infection.

Following equation [1], we fit a series of catalytic models with variable number and duration of age classes to the cumulative incidence of reported cases of RSV by age in Antananarivo across the study period, then compared model fit to the data by AIC to evaluate the most appropriate number and distribution of age classes by which to segregate FOI (Table S2). We first evaluated a null model assuming a single constant FOI across all age classes, then tested increasingly complex models that specified disparate piece-wise constant values for *λ*(*a*) across age brackets (27 models tested in total; Table S2). We focused estimation of heterogeneity in *λ*(*a*) on young and old patients presumed to be at heightened risk for RSV infection. Only models that achieved convergence in the fitting process were evaluated. All models were fit using a quasi-Newton (L-BFGS-B) optimization method in the ‘optim’ function of the base R (v 4.2.2) package ‘stats’; 95% confidence intervals on the resulting age-specific FOI estimates were constructed from the hessian matrix.

#### Seasonal transmission dynamics of RSV via time series Susceptible-Infected-Recovered (TSIR) modeling

We next sought to quantify the seasonality of RSV transmission across our study period, using a time series Susceptible-Infected-Recovered (TSIR) modeling approach implemented in the R package, tsiR (Finkenstädt *et al*. 2000, Bjørnstad *et al*. 2002, Grenfell *et al*. 2002, Becker *et al*. 2017). One of the most widely-used classes of the general SIR model, the TSIR model was originally developed to simplify the process of parameter estimation in fitting SIR models to time series data for measles, a perfectly immunizing, widespread childhood disease (Finkenstädt *et al*. 2000, Bjørnstad *et al*. 2002, Grenfell *et al*. 2002). More recently, TSIR has been adopted for application to other immunizing childhood diseases, including varicella (Baker *et al*. 2018), rubella (Metcalf *et al*. 2011, Wesolowski *et al*. 2016), and RSV (Baker *et al*. 2019, Wambua *et al*. 2022). TSIR depends on two critical assumptions, requiring (*i*), that over long time horizons, the sum of infected cases should be equivalent to the sum of births for highly infectious childhood diseases (for which all individuals are assumed to be eventually exposed), and (*ii*), that the infectious period of the pathogen modeled is equal to the sampling interval for the time series data. TSIR thus captures epidemic dynamics in a series of simple difference equations, whereby the susceptible population in a future timestep (*S*_t+1_) can be modeled as:

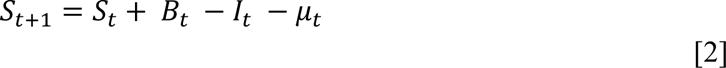

where *B*_t_ corresponds to birth inputs to the population in each timestep, *I*_t_ indicates susceptibles lost to infection, μ_t_ is additive noise, and *S*_t_ can be rewritten as 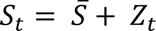 where 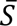 is the mean number of susceptibles in the population and *Z*_t_ describes the (unknown) time-varying deviations around this mean. From this, the susceptible population can be reconstructed iteratively across a time series, after rewriting equation [2] in terms of *Z*_t_ and the starting condition, *Z*_0_:

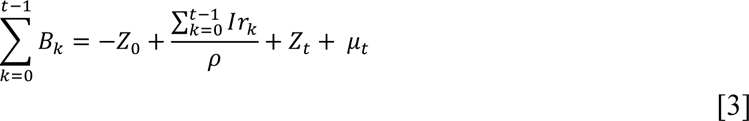

Equation [3] assumes that ρ is the reporting rate of infection and *I*r_k_ is the reported incidence (in our case corresponding to reported cases of RSV in our study region). Because TSIR assumes no overlapping infection generations, the number of infections per timestep can then be expressed as the product of the susceptible and infected populations in the preceding generation, along with the time-varying transmission rate (β_t_):

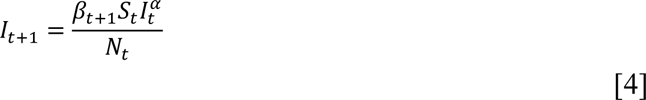

where the homogeneity parameter (*α*) corrects for epidemic saturation in the process of discretizing a continuous epidemic. Equation [4] can then be easily log-linearized as:

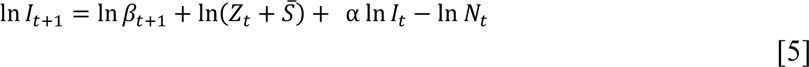

allowing for estimation of β_t_ via simple regression techniques.

From above, we first followed (Baker *et al*. 2019) to reconstruct the population susceptible to RSV in our catchment area in Antananarivo, Madagascar. To this end, we binned RSV positive cases by week across the time series for all reporting sites, and correspondingly summed the population catchments for each site over each week for which data were reported (e.g. the population modeled was higher for timesteps reporting cases from a greater number of sites). To avoid errors in TSIR, we substituted a value of 1 for weeks reporting zero cases. We estimated weekly total births for our study population across the time series by multiplying per capita publicly available birth rates for Madagascar reported by the World Bank (Bank 2023) against the summed population size of the catchment of all reporting study sites, divided evenly across weekly timesteps in a given year (per capita World Bank birth rates are reported as annual means only). Again following (Baker *et al*. 2019), we reconstructed the susceptible population using a simple linear regression of cumulative cases against cumulative births (Table S3). We validated our susceptible reconstruction by comparing the coarse estimate of the basic reproduction number (R_0_) for RSV in Antananarivo that can be derived from the relationship *S* = 1/R_0_ (Wesolowski *et al*. 2016, Becker *et al*. 2017) with that inferred from the mean birth rate and average age of infection across the time series, assuming R_0_ = G/A where G is the inverse of the population birth rate and A is the average age of infection (Rm 1991).

Finally, following susceptible reconstruction, we implemented a log-link linear regression in the ‘Poisson’ family in the tsiR package to estimate the weekly RSV transmission rate (β). Though we modeled our data in 52 weekly timesteps per year, to avoid overfitting, we constrained our fitting to 26 distinct biweekly transmission rates across the 11-year dataset (e.g. transmission was held constant between consecutive two-week intervals; Table S3). Consistent with prior studies (Glass *et al*. 2003, Baker *et al*. 2019), we fixed the homogeneity parameter (α) at 0.97 in the fitting process; previous work has demonstrated that inference into seasonal transmission rates from TSIR is robust to the value of α (Metcalf *et al*. 2009).

#### Climatic drivers of RSV transmission using generalized linear models (GLMs)

Following estimation of seasonal transmission rates for RSV in Antananarivo, we sought to quantify the impact of local climate variables on RSV transmission across the time series. Here, we used a generalized linear model (GLM) in the ‘gaussian family’, fit to the response variable of the natural log of the weekly RSV transmission rate, as estimated from TSIR, with fixed predictor variables of total weekly precipitation (mm), weekly mean relative humidity (%), and mean weekly temperature (^0^C) for Antananarivo across the study period. Our most comprehensive, global GLM thus took the following general form:

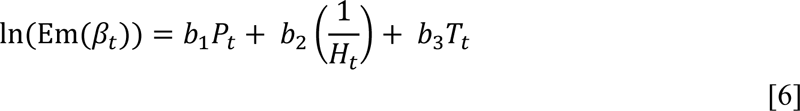

where Em(β_t_) corresponds to the empirically estimated transmission rate from TSIR, and *P*_t_, *H*_t_, and *T*_t_ correspond respectively to total weekly precipitation, mean weekly relative humidity, and mean weekly temperature at time t. Using the ‘dredge’ function in the R package MuMIn (Barton *et al*. 2015), we then conducted model selection by comparing the relative AICc of all predictor variable combinations of the global model defined in equation [6] (Table S4). We used the fitted correlation coefficients of the top-performing model from this selection exercise to explore the predicted impact of climatic variation on RSV transmission. Initially, we also considered GLMs exploring lagged interactions between climate variables and transmission response; however, likely because RSV is a directly-transmitted infection, these representations were not well-supported by the data, and we eventually disregarded this approach.

#### Interannual trends in Antananarivo climate variables and RSV case counts from GAMs

Finally, we aimed to quantify interannual trends for all Antananarivo climate variables considered across the 2011-2021 time series, while controlling for intra-annual seasonal variation. To this end, we fit three separate climate GAMs in the ‘gaussian’ family to the response variables of, respectively, total weekly precipitation (mm), mean weekly relative humidity (%), and mean weekly temperature (^0^C) for Antananarivo. Each GAM incorporated a fixed, numerical effect of ‘year’ and cubic smoothing spline of ‘day of year’ (Table S5). We additionally fit a fourth GAM in the ‘Poisson’ family, which took the same form as the climate GAMs but included a response variable of total weekly reported RSV cases across the time series (Table S5). Subsequently, we restructured all four GAMs with ‘year’ input as a smoothing spline and formatted as factorial, random effect to identify any deviant years in the overall climate or case time series (Table S5).

## RESULTS

### Correlates of RSV infection by GAMs

From January 2011 to December 2021, a total of 3432 samples from reporting sites were screened for RSV. Of these, 989 (28.80%) tested positive for RSV infection (Fig. S1). RSV prevalence varied considerably across the weekly time series, consistent with irregular reporting and seasonality (Fig. 1A).

**Figure 1.**
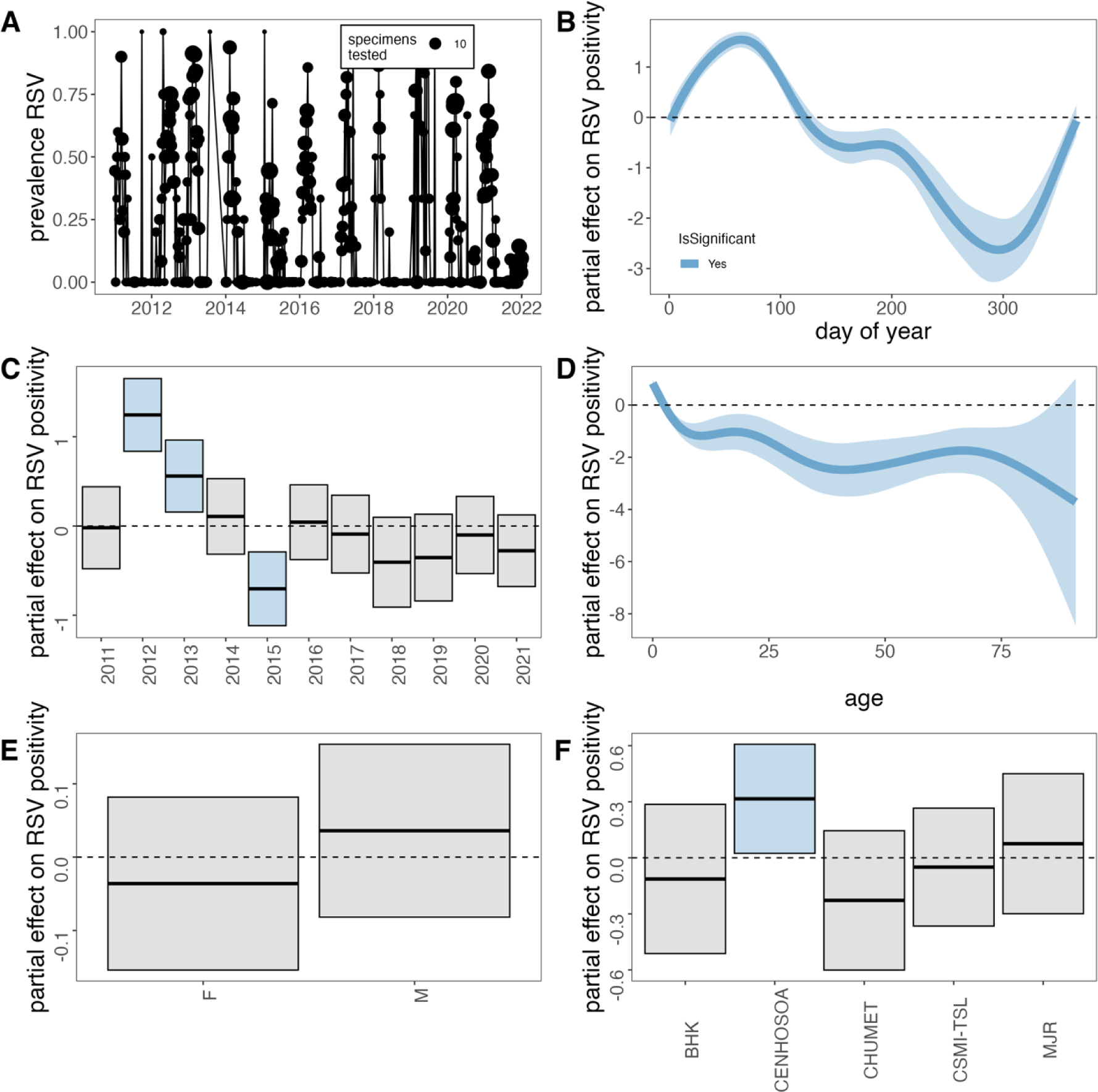
GAM correlates of RSV positivity from influenza surveillance network testing in Antananarivo, Madagascar (2011-2021). (**A**) Time series of weekly RSV prevalence, where point size corresponds to the number of samples tested (total 3432) from January 2011 – December 2021. For GAM fitted to data with all available correlates (3242, partial effect sizes are shown for (**B**) day of year, (**C**) year, (**D**) age, (**E**) sex, and (**F**) sampling locality on the probability of a sample testing positive for RSV infection. All predictors except sex contributed significantly to overall model performance. Shading corresponds to 95% confidence intervals (CI) by standard error. An effect is shaded in gray if the 95% CI crosses zero across the entire range of the predictor variable; in contrast, an effect is shaded in blue and considered “significant” if the 95% CI does not cross zero. Metrics from GAM fits are reported in Table S1.

We first investigated correlates of RSV positivity using GAMs fit to 3242 datapoints for which all possible metadata variables were reported. Our first GAM, in which we modeled year as a factorial random effect performed slightly better than the second GAM modeling year as a numerical variable (Table S1); thus, we report the results of the best fit model only here. In this best fit GAM, we identified a significant effect of day of year, year, age, and reporting sites on RSV positivity; no significant association was identified for sex (Fig. 1; Table S1). In general, days 9-113 of each year (early January through the late April) were significantly positively associated with RSV infection, consistent with records of the peak epidemic season for RSV in Madagascar (Fig. 1B). This relationship turned negative for the rest of the year, with a slight uptick at the end of December, as the onset of a new epidemic season approached. A few discrete years in the time series demonstrated significant deviations in reported RSV prevalence across the time series: years 2012 and 2013 were significantly positively associated with RSV infection, while year 2015 was negatively associated with RSV infection. Patient ages ≤2 years were additionally positively associated with RSV infection. Most other ages were negatively associated with RSV infection, though no significant directionality in association was identified in patient ages >85 years (Fig. 1D). The reporting hospital of CENHOSOA was also positively associated with RSV infection (Fig. 1F), likely a result of the overwhelming predominance of samples received from this locality. Day of year and annual patterns were largely recapitulated when modeling (in a third GAM) the entire dataset of 3432 samples without including age, sex, and reporting hospital as predictors: in this model, days of year 7-111 were significantly positively associated with RSV infection, as were discrete years 2012, 2013, and 2019. As in the first GAM, year 2015 was also significantly negatively associated with RSV positivity (Table S1).

### Estimation of the age-structured FOI for RSV cases

Of the catalytic models tested, a model incorporating a piecewise, age-specific FOI across nine discrete age bins (with respective lower bounds of 0, 0.5, 1, 2, 3, 4, 25, 60, and 70 years) offered the best fit to the data (Table S2). Within this model framework, we estimated the highest FOIs in the two youngest cohorts (FOI = 1.03 [0.92-1.13] for ages 0 to <0.5 years; FOI = 1.50 [1.27-l.73] for ages 0.5 to <1 year) (Fig. 2). From this peak, FOI then decreased with increasing age across the first four years of life and sank to near-zero between ages 4 – 60 years (Fig. 2). FOI increased again in individuals aged 60 to <70 years (0.11 [0.05-0.16]) and individuals 70+ years in age (0.34 [0.15-0.51]). In general, we found improved model fit to the data when incorporating heightened specificity in *λ*(*a*) for CU5, as well as in individuals >60 years in age. Additional complexity in modeled age structure in older adolescents or young-and middle-aged adults did little to improve model performance (Table S2). Consistent with the literature, these patterns suggest that the highest hazard of RSV infection is concentrated in the youngest and, to a lesser extent, oldest populations in Antananarivo.

**Figure 2.**
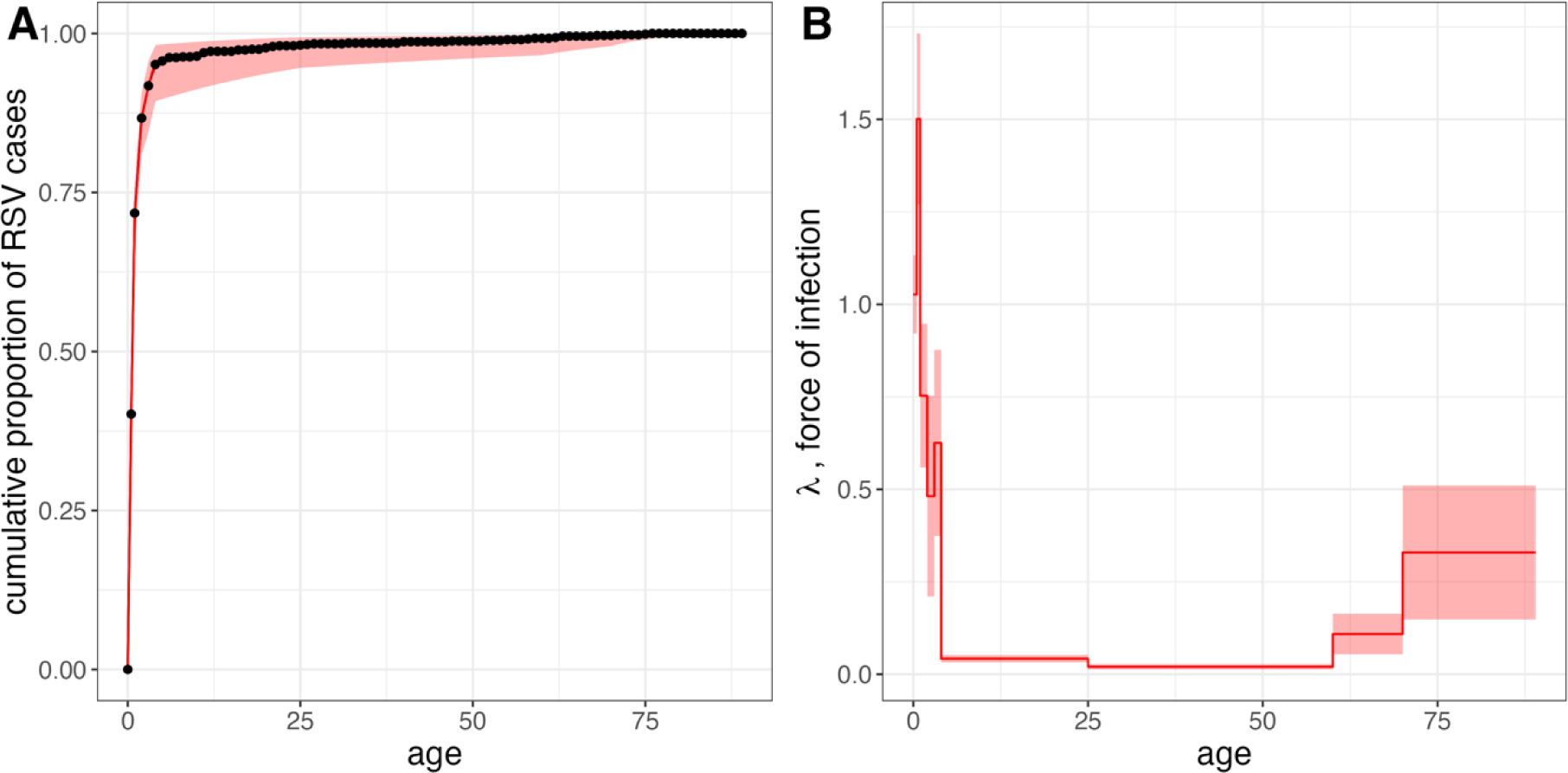
Catalytic model estimation of age-structured force of infection for RSV hospitalizations. (**A**) Cumulative proportion of hospitalized cases by age, across 2011-2021 study period from all focal hospitals in Antananarivo (total = 3259 datapoints). Black dots and connecting black line correspond to raw data binned across annual age brackets, with the first year of life separated into two age brackets (<0.5 years and ≥0.5 years). Red line corresponds to model output from best-fit FOI model, incorporating 9 piecewise constant, age-specific hazards of infection across discrete age classes, as shown in (**B**). In both panels, red shading corresponds to 95% CI on FOI estimates computed from the hessian matrix during model fitting; upper and lower CI bounds are used to project 95% CIs on modeled cumulative incidence in panel A. Comparisons of all models tested and exact FOI estimates from best-fit model are reported in Table S2.

### Seasonality of RSV transmission by TSIR modeling

We next binned reported RSV cases into weekly intervals across our time series for TSIR modeling. Consistent with results from the binomial GAMs, we observed from the raw data that weekly cases were highest in the first third of each year (January – April), excepting the year 2012, for which the epidemic peak was observed between May and August (Fig. 3A). Our efforts to reconstruct the population susceptible to RSV from publicly available birth rate data indicated significant underreporting of RSV across the time series, with a mean estimated reporting rate of only 2.9% (Fig. S2A-D). Nonetheless, the regression of cumulative reported cases against cumulative births was well-supported (R^2^= 0.98; Table S3) and produced a plausible estimate for the mean susceptible population of roughly 20% (Fig. S2D). This quantity corresponds to an estimated R_0_ = 5 for RSV in our Antananarivo catchment area; coarse estimation of R_0_ using the inverse of the mean per capita birth rate (0.0327) and average age of infection (6.67) across the time series yields a comparable estimate of R_0_ = 4.58. Literature reported R_0_ values for RSV range from 1–9, with 3 as the most commonly-cited statistic (Weber *^e^t al.* 2001, White *et al*. 2007, Pitzer *et al*. 2015, Reis *et al*. 2016, Reis *et al*. 2018). Though our estimates are in the upper half of this range, R_0_ = 4.5–5 for RSV is not illogical given high birth rates and densely aggregated populations in Antananarivo.

**Figure 3.**
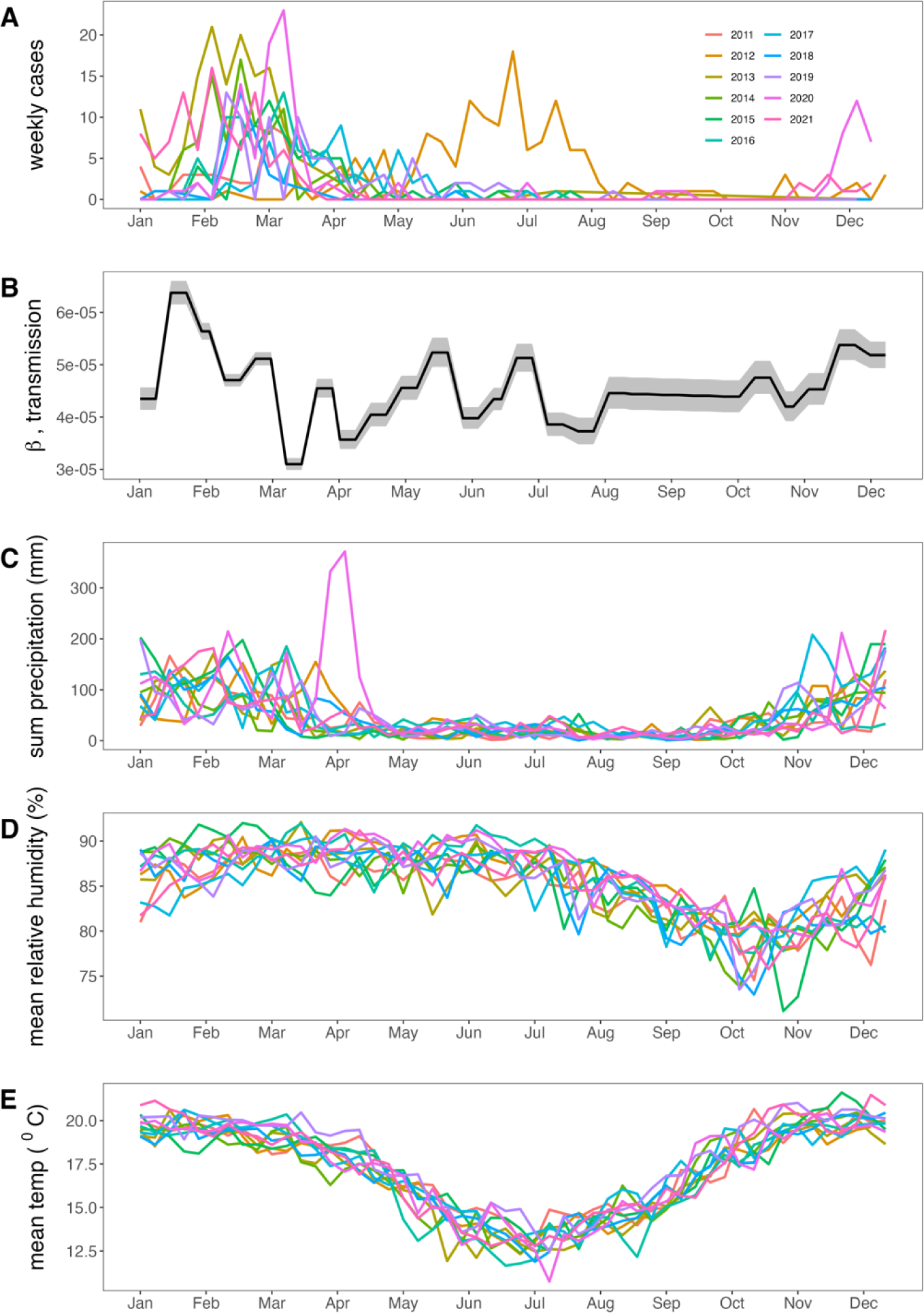
Seasonal trends in RSV infection and climate variables for Antananarivo, Madagascar (2011 – 2021). (**A**) Weekly total RSV cases across all reporting hospitals in our catchment area, from January 2011 – December 2021, colored by year of reporting as indicated in legend. (**B**) Weekly RSV transmission rate, as estimated from TSIR model fitting across the year; shading corresponds to 95% confidence intervals by standard error from GLM fit. Raw weekly climate data for (**C**) total precipitation (mm), (**D**) mean relative humidity (%), and (**E**) mean temperature (^0^C) in Antananarivo across the time series, colored by year of reporting as indicated in panel **A**.

Consistent with prior patterns observed in the GAMs (Fig. 1) and in the raw data (Fig. 3A), our fitted TSIR model estimated that RSV transmission peaked in January–February, slightly preceding cases, then declined throughout the rest of the year before climbing again in December (Fig. 3B; Fig S2E). As with susceptible reconstruction, our log-link linear regression to estimate transmission was well supported (McFadden pseudo-R^2^= 0.64; Table S3) and successfully reconstructed the time series of cases following simulation (Fig. S2F-G).

### Climate predictors of seasonal RSV transmission by GLMs

From the Antananarivo raw climate data, we observed that annual precipitation largely mirrored the seasonality of RSV, with most of the rainfall concentrated between January and the end of April and a slight increase in November and December leading up to the onset of each new year (Fig. 3C). Relative humidity also peaked in the early half of the year but remained high through June, with lows concentrated between September and November (Fig. 3D). Mean temperature followed the same broad pattern as the other climate variables but with a more gradual, sinusoidal annual cycle, with highs concentrated in the summer between November – March and lows between June – August (Fig. 3E).

Our top performing GLM to evaluate the strength of association between climate and RSV transmission (as estimated from TSIR) included all three weekly Antananarivo climate variables (precipitation, humidity, and temperature) as significant correlates of transmission, with precipitation contributing the most to the total deviance explained by the model (R^2^=0.10; Fig. 4A-B; Table S4). The second-best performing model included only precipitation and humidity as significant correlates of RSV transmission, again with a more pronounced effect for precipitation. In all cases, precipitation demonstrated a highly significant, positive correlation with the weekly RSV transmission rate (p<0.001***; Fig. 4B-C; Table S4). By contrast, relative humidity was significantly negatively associated with RSV transmission (p<0.01*; Fig. 4B-C; Table S4). Mean weekly temperature was positively correlated with transmission, but the association was considerably weaker than that observed for precipitation and only marginally significant (p<0.1*; Fig. 4B-C; Table S4).

**Figure 4.**
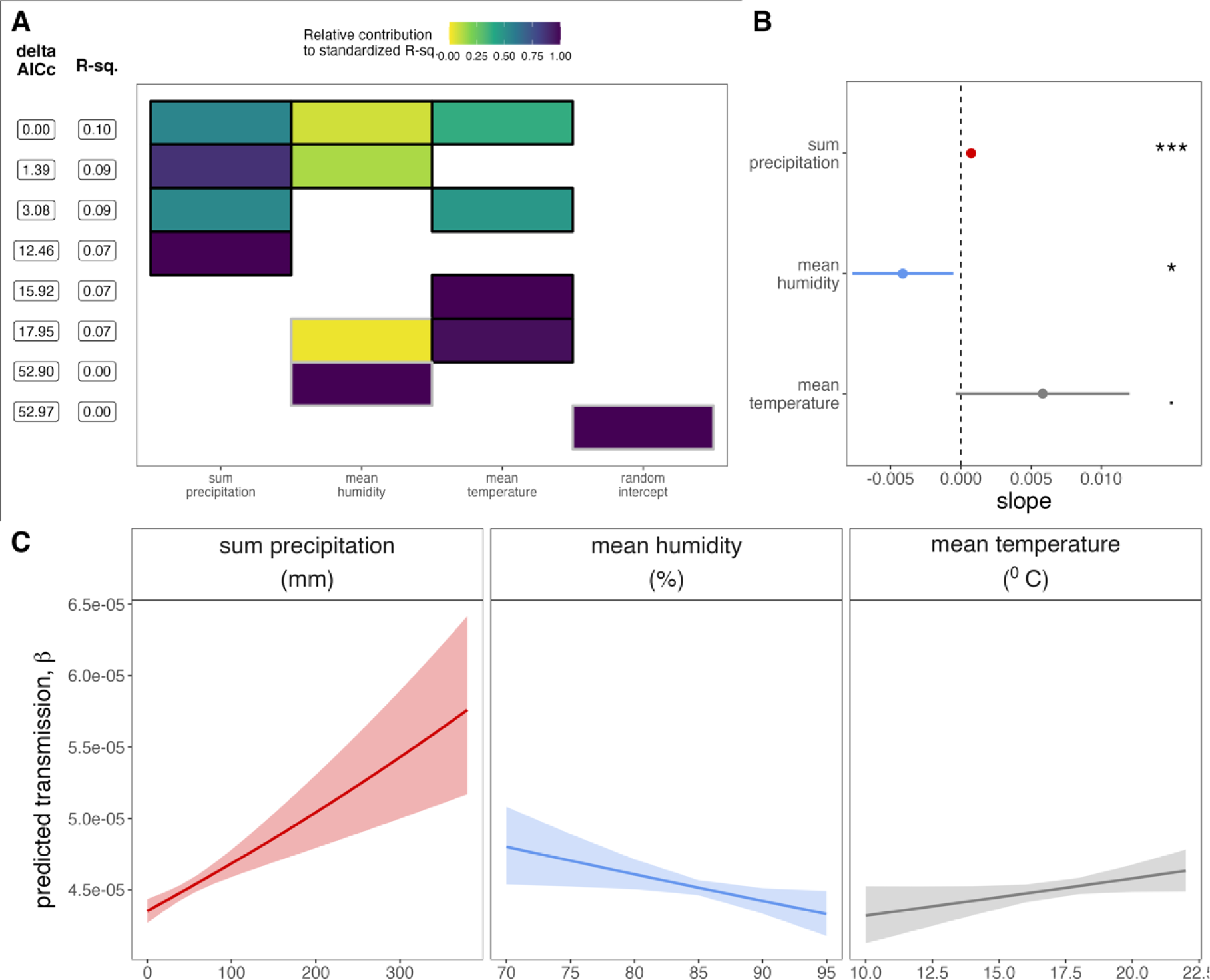
Climatic correlates of RSV transmission, from GLM analysis. (**A**) Top 8 GLMs using climate variables to predict RSV transmission from TSIR, ranked by δAICc. Rows represent individual models, and columns represent predictor variables. Cells are shaded according to the proportion of deviance explained by each predictor. Cells representing predictor variables with a p-value significance level of <0.1 are outlined in black; all others are outlined in gray. (**B**) Effect size of significant climate correlates from best-fit GLM (top row in panel **A**) on response variable of RSV transmission. 95% confidence intervals by standard error are shown as horizontal bars and corresponding p-values are highlighted to the right for: precipitation (p<0.001***), humidity (p<0.05*), and temperature (p<.1). Statistical output is reported in Table S4. (**C**) Predicted response of RSV transmission to a range of values for significant climate correlates from best-fit GLM (top row in panel **A**); shading corresponds to 95% confidence intervals by standard error, computed for each effect size.

### Interannual trends for climate variables and RSV cases by GAMs

GAMs fit to the 2011-2021 time series for all three Antananarivo climate variables demonstrated a significant, positive interannual trend in all cases, suggesting that precipitation, humidity, and temperature have all increased across the past decade (precipitation: p=0.05*; humidity: p=0.07*; temperature: p=0.004**; Fig. S3 A-C; Table S5). Intra-annual trends captured in GAMs mimicked those previously described from observations of the raw data, with precipitation peaking at the beginning and end of each calendar year, relative humidity offset but still at its highest in the early half of the year, and temperature more evenly distributed with highs concentrated between November – March and lows from June – August (Fig. S3A-C; Fig. 3C-E). In contrast to climate patterns, a Poisson GAM fit to the weekly time series of case counts demonstrated no significant interannual trend (p>0.1; Fig S3D; Table S5), after controlling for intra-annual cycling. Climate GAMs considering year as a factorial random effect indicated that year 2020 experienced significantly higher precipitation and humidity than average, while 2019 was found to be significantly warmer (Fig. S3); by contrast, year 2011 was found to have lower-than-average humidity and years 2013 and 2016 to be significantly cooler than average (Fig. S3). We identified no correspondence between those years exhibiting anomalous climate behavior and those previously identified to have a higher-than-average RSV burden (2012 and 2013); RSV deviations already explored in Fig. 1 were largely confirmed by the Poisson GAM tested here (Fig. S3).

## DISCUSSION

In this study, we used a suite of modeling approaches to (1) characterize correlates for reported RSV infections, (2) quantify age-structure in the force of RSV infection, (3) document seasonality in RSV transmission, and (4) identify possible climatic drivers of this seasonality in the low income setting of Antananarivo, Madagascar.

The results of our analysis of correlates of RSV transmission are consistent with those previously documented for this disease, both in Madagascar and elsewhere (Hall *et al*. 2013, Razanajatovo *et al*. 2018, Razanajatovo *et al*. 2022): we find that reported hospitalizations for RSV are largely concentrated in very young patients (≤ 2 years of age) with a potential weaker association in elderly individuals (>60 years of age). Our quantification of the age-structured FOI for RSV indicates that the most intense transmission is focused in infants aged less than one year, with transmission intensity steadily decreasing to near-zero after four years of age, before rising again in elderly cohorts. Because we fit our FOI models to reported cases only, it is possible that less pathogenic RSV transmission may dominate among older children, resulting in asymptomatic infections, as has been previously suggested in the analysis of age-structured *serological* data from other regions (Nakajo *et al*. 2023). Our crude estimates for RSV R_0_, which we computed from the average age of infection and mean population-level birth rate, as well as from the mean susceptible population estimated from TSIR, additionally suggest that RSV transmission is elevated in Antananarivo, as compared with global averages (Weber *et al*. 2001, White *et al*. 2007, Pitzer *et al*. 2015, Reis *et al*. 2016, Reis *et al*. 2018). Again, our reliance on reported cases likely upwardly biases these calculations by overrepresenting more pathogenic strains or more vulnerable populations or both. Nonetheless, these estimates fall well within reported ranges for RSV R_0_ (Weber *et al*. 2001, White *et al*. 2007, Pitzer *et al*. 2015, ^R^eis *et al*. 201^6^, Reis *et al*. 2018), and a slightly elevated RSV transmission rate is not illogical, given Antananarivo’s young and densely aggregated population.

In addition to describing the magnitude and age-structure of RSV transmission, our study also successfully quantifies intra-annual seasonality in RSV dynamics in Antananarivo. A seasonal concentration of RSV burden in the first half of the calendar year in Madagascar has been previously suggested based on more qualitative examination of the raw case data (Rabarison *et al*. 2019, Razanajatovo *et al*. 2022); however, our study is the first to actually quantify RSV transmission, which slightly precedes cases. Our investigation of climatic correlates of the weekly transmission rate echoes recent work from the northern hemisphere which highlights a significant positive role for precipitation and negative role for humidity in driving RSV transmission dynamics (Baker *et al*. 2019). In addition, we identify a significant (though weaker) positive correlation between RSV transmission and temperature in Antananarivo. Because high temperatures and high precipitation are themselves correlated in tropical Madagascar’s rainy season, which spans October – March, it is possible that this muted contribution of temperature to RSV transmission dynamics may simply be an artifact of the precipitation driver, rather than a causative association of its own. Our examination of interannual trends in the climate data suggests that precipitation, humidity, and temperature have all increased across the past decade study period in Antananarivo, but that average RSV caseload has remained constant. As high precipitation is associated with elevated RSV transmission rates, but high humidity is associated with lower RSV transmission rates in our dataset, it is possible that these climate drivers have had largely neutralizing effects on the overall RSV burden throughout our time series. Nonetheless, our statistical models demonstrate a more pronounced effect of precipitation on the overall variation in observed RSV transmission, as has been reported in other tropical locations that demonstrate high average annual humidity with minimal variation across the year (Baker *et al*. 2019). Though there is no animal model available for RSV, experimental work in influenza has demonstrated that low humidity conditions favor transmission between guinea pigs, as a result of either increased survival of the virus and/or extended duration of virus circulation in aerosols in drier conditions (Lowen et al. 2007). Future climate impacts on RSV transmission are likely to depend on the relative slope of each climate driver’s projected increase in a specific locality: over the past decade, precipitation has increased at a faster rate than humidity in tropical Antananarivo. If current trends hold, higher precipitation could drive future RSV transmission beyond the tempering effects of humidity.

Finally, our analysis highlights a few years in our decade-long time series that vary significantly from the overall trend, with higher-or lower-than-average RSV-attributed infections and, in one year (2012), an irregular, off-season peak. Prior work in other systems has described off-season RSV epidemics attributed to irregular climate patterns (De Silva *et al*. 1986), but we were unable to identify any association between deviant years in the RSV time series in Antananarivo and any observed climatic anomalies. Higher-or lower-than average RSV burden in a given year could instead reflect the complex interplay between host immunity and circulating viral genotypes—both those of RSV itself, for which two major subtypes are known (Peret *et al*. 1998), and those of other respiratory infections known to induce some degree of anti-RSV cross-protective immunity (Bhattacharyya *et al*. 2015). Prior sequencing efforts of Antananarivo RSV cases over the same time period indicate that year 2012 witnessed a transition in caseload from the previously dominant RSV subtype B to RSV subtype A – genotype NA1, which was subsequently replaced by the introduction of the novel RSV subtype A – genotype ON1 in 2014 (Eshaghi *et al*. 2012, Razanajatovo Rahombanjanahary *et al*. 2020)). It is possible that the irregular seasonality of the RSV epidemic of 2012 reflects a lack of prior immunity in the Antananarivo CU5 population to RSV subtype B prior to turnover—though this hypothesis is impossible to test in the absence of additional viral genomic sequencing and paired subtype-specific serology. Off-season RSV transmission was well-documented globally in 2020 and 2021 following relaxation of non-pharmaceutical interventions (NPIs) implemented to counter the initial spread of SARS-CoV-2 (Eden *et al*. 2022). Intriguingly, our analysis did not recover any signature of aberrant RSV transmission for the year 2020 or 2021 in Antananarivo, suggesting that NPIs were largely unsuccessful at reducing respiratory virus transmission in this region during this time (Razanajatovo *et al*. 2022). Indeed, broadscale serosurveys in 2020 and 2021 suggest that over 40% of the Antananarivo population was exposed to SARS-CoV-2 within the first six months of the pandemic (Razafimahatratra *et al*. 2021), underscoring the relative ineffectiveness of NPIs on reducing respiratory disease burden. Our study is limited by its focus to only a few reporting sites in Antananarivo, with most samples received from one sentinel site (CENHOSOA). This restricted geography somewhat impedes our ability to draw conclusions about broad trends in RSV dynamics across Madagascar. In addition, our FOI estimation is limited by the necessarily arbitrary segregation of the population into discrete age classes; while we attempted to consider all plausibly relevant delineations of age with respect to transmission, it is possible that we may have overlooked important dynamics hidden in untested hypotheses. In addition, we were challenged by uncertainty in estimates of the population served by the catchment area of focal sites, which subsequently undermines downstream estimation of local birth rates and corresponding susceptible population reconstruction for input into TSIR. As a frequent limitation of TSIR approaches, we were forced to extrapolate weekly birth rate data for our study population using very broad national-level, annual birth rate estimates from the World Bank. More fine scale birth data specifically tailored to our study region would do much to improve inference into infection dynamics. Our climate records, which draw from Antananarivo at large, are similarly broad, given the highly localized (e.g. within-household) nature of the majority of RSV transmission (Hall *et al*. 1976, Munywoki *et al*. 2014). Finally, as mentioned, our dataset was limited to reported RSV cases only. More widespread prospective sampling of less virulent disease manifestations – either through molecular testing, or more feasibly, serology – would greatly enhance our inference into RSV transmission. In addition, investigation of genome sequences would enable us to test hypotheses about the impact of virus genotype diversity on asynchronous and off-season dynamics. Despite these challenges, our modeling efforts recover plausible estimates for both the seasonality and R_0_ of reported cases of RSV in Antananarivo, suggesting that these uncertainties did not seriously impact our results.

All told, our study underscores the heavy morbidity and mortality burden that RSV presents to young children in Antananarivo and highlights an important role for climate in driving seasonal epidemics. Future changes in climatic parameters, particularly precipitation, are likely to impact RSV dynamics and may impact transmission, at least in the short-term, in Madagascar. Introductions of new intervention strategies are greatly needed to mitigate RSV’s heavy mortality burden in LMICs—especially considering possible intensification of burden in response to climate. RSV vaccines for older patients and pregnant mothers, as well as monoclonal antibody treatment for neonates, have recently become available in high income countries (Langedijk *et al*. 2023). Despite projections of positive impact (Brand *et al*. 2020), these interventions have not yet reached the Global South, largely as a result of high cost barriers and lack of awareness of regional health authorities and communities regarding the burden of RSV. Shortages of human, financial, and material resources, which jeopardize the provisioning of quality health services, are still a serious challenge in LMICs. Equitable global distribution of these interventions needs to be a major public health priority for the next decade.

## Data Availability

All data produced in the present study are available upon reasonable request to the authors

https://github.com/brooklabteam/RSV-madagascar

https://power.larc.nasa.gov/data-access-viewer/

## ACKNOWLEDGEMENTS

The authors thank all clinicians and nurses involved in the study, as well as staff involved in daily surveillance reporting of ILI and SARI cases at sentinel sites. The authors additionally thank the laboratory technicians at the Virology Unit of IPM and the Clinical and Medical Laboratory for their tremendous work in the molecular testing of the biological samples.

## CONTRIBUTORS

CEB and JMH conceived the project and acquired the funding, in collaboration with NRS, and VL. Influenza and RSV Sentinel surveillance in Madagascar was coordinated by JMH, LR, NHR, and JHR. Sample collection was carried out by LR, JHR, and AR. All laboratory work, involving RNA extraction and subsequent PCR, was conducted by THR, NHR, and CR. CEB led the statistical analysis and FOI and TSIR modeling, with support from HCR and THR. THR and CEB co-wrote the original draft of the manuscript. All authors including DADR edited and approved the manuscript.

## ETHICS APPROVAL

This study was approved by the Ethics Committee of the Ministry of Health (MoH) of Madagascar (number: 96-MSANP/CERBM, dated August 27, 2018).

## REFERENCES

1. Andeweg SP, Schepp RM, van de Kassteele J, Mollema L, Berbers GA and van Boven M (2021). Population-based serology reveals risk factors for RSV infection in children younger than 5 years. Scientific Reports 11(1): 8953.

2. Baker RE, Mahmud AS and Metcalf CJE (2018). Dynamic response of airborne infections to climate change: predictions for varicella. Climatic change 148: 547–560.

3. Baker RE, Mahmud AS, Wagner CE, Yang W, Pitzer VE, Viboud C, Vecchi GA, Metcalf CJE and Grenfell BT (2019). Epidemic dynamics of respiratory syncytial virus in current and future climates. Nature communications 10(1): 5512.

4. Bank TW (2023). The World Bank in Madagascar.

5. Barton K and Barton MK (2015). Package ‘mumin’. Version 1(18): 439.

6. Becker AD and Grenfell BT (2017). tsiR: An R package for time-series Susceptible-Infected-Recovered models of epidemics. PLoS One 12(9): e0185528.

7. Bhattacharyya S, Gesteland PH, Korgenski K, Bjørnstad ON and Adler FR (2015). Cross-immunity between strains explains the dynamical pattern of paramyxoviruses. Proceedings of the National Academy of Sciences 112(43): 13396–13400.

8. Bjørnstad ON, Finkenstädt BF and Grenfell BT (2002). Dynamics of measles epidemics: estimating scaling of transmission rates using a time series SIR model. Ecological monographs 72(2): 169–184.

9. Brand SP, Munywoki P, Walumbe D, Keeling MJ and Nokes DJ (2020). Reducing respiratory syncytial virus (RSV) hospitalization in a lower-income country by vaccinating mothers-to-be and their households. Elife 9: e47003.

10. Chew FT, Doraisingham S, Kumarasinghe G, Lee BW and Ling AE (1998). Seasonal trends of viral respiratory tract infections in the tropics. Epidemiology and Infection 121(1): 121–128.

11. Data OWi (2019). Causes of death in children under five, World, 2019.

12. De Silva L and Hanlon M (1986). Respiratory syncytial virus: Report of a 5-year study at a children’s hospital. Journal of medical virology 19(4): 299–305.

13. Desa U (2018). The 2018 Revision of World Urbanization Prospects. New York: UN DESA.

14. Eden J-S, Sikazwe C, Xie R, Deng Y-M, Sullivan SG, Michie A, Levy A, Cutmore E, Blyth CC and Britton PN (2022). Off-season RSV epidemics in Australia after easing of COVID-19 restrictions. Nature communications 13(1): 2884.

15. Eshaghi A, Duvvuri VR, Lai R, Nadarajah JT, Li A, Patel SN, Low DE and Gubbay JB (2012). Genetic variability of human respiratory syncytial virus A strains circulating in Ontario: a novel genotype with a 72 nucleotide G gene duplication. PLoS One 7(3): e32807.

16. Falsey AR, Hennessey PA, Formica MA, Cox C and Walsh EE (2005). Respiratory syncytial virus infection in elderly and high-risk adults. New England Journal of Medicine 352(17): 1749–1759.

17. Finkenstädt BF and Grenfell BT (2000). Time series modelling of childhood diseases: a dynamical systems approach. Journal of the Royal Statistical Society Series C: Applied Statistics 49(2): 187–205.

18. Glass K, Xia Y and Grenfell BT (2003). Interpreting time-series analyses for continuous-time biological models—measles as a case study. Journal of theoretical biology 223(1): 19–25.

19. Grenfell BT and Anderson R (1985). The estimation of age-related rates of infection from case notifications and serological data. Epidemiology & Infection 95(2): 419–436.

20. Grenfell BT, Bjørnstad ON and Finkenstädt BF (2002). Dynamics of measles epidemics: scaling noise, determinism, and predictability with the TSIR model. Ecological monographs 72(2): 185–202.

21. Haber N (2018). Respiratory syncytial virus infection in elderly adults. Med Mal Infect 48(6): 377–382.

22. Hall CB, Geiman JM, Biggar R, Kotok DI, Hogan PM and Douglas Jr RG (1976). Respiratory syncytial virus infections within families. New England Journal of Medicine 294(8): 414–419.

23. Hall CB, Weinberg GA, Blumkin AK, Edwards KM, Staat MA, Schultz AF, Poehling KA, Szilagyi PG, Griffin MR, Williams JV, Zhu Y, Grijalva CG, Prill MM and Iwane MK (2013). Respiratory syncytial virus-associated hospitalizations among children less than 24 months of age. Pediatrics 132(2): e341–348.

24. Heisey DM, Joly DO and Messier F (2006). The fitting of general force-of-infection models to wildlife disease prevalence data. Ecology 87(9): 2356–2365.

25. Heraud JM, Razanajatovo NH and Viboud C (2019). Global circulation of respiratory viruses: from local observations to global predictions. Lancet Glob Health 7(8): e982–e983.

26. Hogan AB, Glass K, Moore HC and Anderssen RS (2016). Exploring the dynamics of respiratory syncytial virus (RSV) transmission in children. Theor Popul Biol 110: 78–85.

27. Langedijk AC and Bont LJ (2023). Respiratory syncytial virus infection and novel interventions. Nature Reviews Microbiology 21(11): 734–749.

28. Li Y, Wang X, Blau DM, Caballero MT, Feikin DR, Gill CJ, Madhi SA, Omer SB, Simões EAF, Campbell H, Pariente AB, Bardach D, Bassat Q, Casalegno JS, Chakhunashvili G, Crawford N, Danilenko D, Do LAH, Echavarria M, Gentile A, Gordon A, Heikkinen T, Huang QS, Jullien S, Krishnan A, Lopez EL, Markić J, Mira-Iglesias A, Moore HC, Moyes J, Mwananyanda L, Nokes DJ, Noordeen F, Obodai E, Palani N, Romero C, Salimi V, Satav A, Seo E, Shchomak Z, Singleton R, Stolyarov K, Stoszek SK, von Gottberg A, Wurzel D, Yoshida LM, Yung CF, Zar HJ and Nair H (2022). Global, regional, and national disease burden estimates of acute lower respiratory infections due to respiratory syncytial virus in children younger than 5 years in 2019: a systematic analysis. Lancet 399(10340): 2047–2064.

29. Linssen RS, Bem RA, Kapitein B, Rengerink KO, Otten MH, den Hollander B, Bont L and van Woensel JBM (2021). Burden of respiratory syncytial virus bronchiolitis on the Dutch pediatric intensive care units. Eur J Pediatr 180(10): 3141–3149.

30. Long GH, Sinha D, Read AF, Pritt S, Kline B, Harvill ET, Hudson PJ and Bjørnstad ON (2010). Identifying the age cohort responsible for transmission in a natural outbreak of Bordetella bronchiseptica. PLoS Pathogens 6(12): e1001224.

31. Lowen AC, Mubareka S, Steel J and Palese P (2007). Influenza virus transmission is dependent on relative humidity and temperature. PLoS Pathogens 3(10): e151.

32. Mathisen M, Strand TA, Sharma BN, Chandyo RK, Valentiner-Branth P, Basnet S, Adhikari RK, Hvidsten D, Shrestha PS and Sommerfelt H (2009). RNA viruses in community-acquired childhood pneumonia in semi-urban Nepal; a cross-sectional study. BMC medicine 7: 1–12.

33. Matthew J, Pinto Pereira LM, Pappas TE, Swenson CA, Grindle KA, Roberg KA, Lemanske RF, Lee W-M and Gern JE (2009). Distribution and seasonality of rhinovirus and other respiratory viruses in a cross-section of asthmatic children in Trinidad, West Indies. Italian Journal of Pediatrics 35(1): 1–10.

34. Metcalf CJE, Bjørnstad O, Ferrari M, Klepac P, Bharti N, Lopez-Gatell H and Grenfell BT (2011). The epidemiology of rubella in Mexico: seasonality, stochasticity and regional variation. Epidemiology & Infection 139(7): 1029–1038.

35. Metcalf CJE, Bjørnstad ON, Grenfell BT and Andreasen V (2009). Seasonality and comparative dynamics of six childhood infections in pre-vaccination Copenhagen. Proceedings of the Royal Society B: Biological Sciences 276(1676): 4111–4118.

36. Muench H (1959). Catalytic models in epidemiology, Harvard University Press.

37. Munywoki PK, Koech DC, Agoti CN, Lewa C, Cane PA, Medley GF and Nokes DJ (2014). The source of respiratory syncytial virus infection in infants: a household cohort study in rural Kenya. The Journal of Infectious Diseases 209(11): 1685–1692.

38. Nair H, Nokes DJ, Gessner BD, Dherani M, Madhi SA, Singleton RJ, O’Brien KL, Roca A, Wright PF and Bruce N (2010). Global burden of acute lower respiratory infections due to respiratory syncytial virus in young children: a systematic review and meta-analysis. The Lancet 375(9725): 1545–1555.

39. Nakajo K and Nishiura H (2023). Age-dependent risk of respiratory syncytial virus infection: A systematic review and hazard modeling from serological data. The Journal of Infectious Diseases: jiad147.

40. NASA National Aeronautics and Space Administration (NASA) database

41. O’Brien KL, Baggett HC, Brooks WA, Feikin DR, Hammitt LL, Higdon MM, Howie SR, Knoll MD, Kotloff KL and Levine OS (2019). Causes of severe pneumonia requiring hospital admission in children without HIV infection from Africa and Asia: the PERCH multi-country case-control study. The Lancet 394(10200): 757–779.

42. Obando-Pacheco P, Justicia-Grande AJ, Rivero-Calle I, Rodríguez-Tenreiro C, Sly P, Ramilo O, Mejías A, Baraldi E, Papadopoulos NG, Nair H, Nunes MC, Kragten-Tabatabaie L, Heikkinen T, Greenough A, Stein RT, Manzoni P, Bont L and Martinón-Torres F (2018). Respiratory Syncytial Virus Seasonality: A Global Overview. The Journal of Infectious Diseases 217(9): 1356–1364.

43. Obolski U, Kassem E, Na’amnih W, Tannous S, Kagan V and Muhsen K (2021). Unnecessary antibiotic treatment of children hospitalised with respiratory syncytial virus (RSV) bronchiolitis: risk factors and prescription patterns. Journal of Global Antimicrobial Resistance 27: 303–308.

44. Peret TC, Hall CB, Schnabel KC, Golub JA and Anderson LJ (1998). Circulation patterns of genetically distinct group A and B strains of human respiratory syncytial virus in a community. J Gen Virol 79 (Pt 9): 2221–2229.

45. Pinquier D, Crépey P, Tissières P, Vabret A, Roze JC, Dubos F, Cahn-Sellem F, Javouhey E, Cohen R and Weil-Olivier C (2023). Preventing Respiratory Syncytial Virus in Children in France: A Narrative Review of the Importance of a Reinforced Partnership Between Parents, Healthcare Professionals, and Public Health Authorities. Infect Dis Ther 12(2): 317–332.

46. Pitzer VE, Viboud C, Alonso WJ, Wilcox T, Metcalf CJ, Steiner CA, Haynes AK and Grenfell BT (2015). Environmental drivers of the spatiotemporal dynamics of respiratory syncytial virus in the United States. PLoS Pathogens 11(1): e1004591.

47. Pomeroy LW, Bjørnstad ON, Kim H, Jumbo SD, Abdoulkadiri S and Garabed R (2015). Serotype-specific transmission and waning immunity of endemic foot-and-mouth disease virus in Cameroon. PLoS One 10(9): e0136642.

48. Rabarison JH, Rakotondramanga JM, Ratovoson R, Masquelier B, Rasoanomenjanahary AM, Dreyfus A, Garchitorena A, Rasambainarivo F, Razanajatovo NH, Andriamandimby SF, Metcalf CJ, Lacoste V, Heraud JM and Dussart P (2023). Excess mortality associated with the COVID-19 pandemic during the 2020 and 2021 waves in Antananarivo, Madagascar. BMJ Glob Health 8(7).

49. Rabarison JH, Tempia S, Harimanana A, Guillebaud J, Razanajatovo NH, Ratsitorahina M and Heraud JM (2019). Burden and epidemiology of influenza-and respiratory syncytial virus-associated severe acute respiratory illness hospitalization in Madagascar, 2011-2016. Influenza and Other Respiratory Viruses 13(2): 138-147.

50. Randrianasolo L, Raoelina Y, Ravololomanana L, Randrianarivo-Solofoniaina A, Héraud J-M and Richard V (2010). Surveillance sentinelle des fièvres à Madagascar. Revue d’Épidémiologie et de Santé Publique 58: S88.

51. Rasolofonirina N (2003). Historique de la grippe à Madagascar. Archives de l’Institut Pasteur de Madagascar 69(1).

52. Razafimahatratra SL, Ndiaye MDB, Rasoloharimanana LT, Dussart P, Sahondranirina PH, Randriamanantany ZA and Schoenhals M (2021). Seroprevalence of ancestral and Beta SARS-CoV-2 antibodies in Malagasy blood donors. The Lancet global health 9(10): e1363–e1364.

53. Razanajatovo NH, Guillebaud J, Harimanana A, Rajatonirina S, Ratsima EH, Andrianirina ZZ, Rakotoariniaina H, Andriatahina T, Orelle A, Ratovoson R, Irinantenaina J, Rakotonanahary DA, Ramparany L, Randrianirina F, Richard V and Heraud JM (2018). Epidemiology of severe acute respiratory infections from hospital-based surveillance in Madagascar, November 2010 to July 2013. PLoS One 13(11): e0205124.

54. Razanajatovo NH, Randriambolamanantsoa TH, Rabarison JH, Randrianasolo L, Ankasitrahana MF, Ratsimbazafy A, Raherinandrasana AH, Razafimanjato H, Raharinosy V, Andriamandimby SF, Heraud JM, Dussart P and Lacoste V (2022). Epidemiological Patterns of Seasonal Respiratory Viruses during the COVID-19 Pandemic in Madagascar, March 2020-May 2022. Viruses 15(1).

55. Razanajatovo NH, Richard V, Hoffmann J, Reynes JM, Razafitrimo GM, Randremanana RV and Heraud JM (2011). Viral etiology of influenza-like illnesses in Antananarivo, Madagascar, July 2008 to June 2009. PLoS One 6(3): e17579.

56. Razanajatovo Rahombanjanahary NH, Rybkina K, Randriambolamanantsoa TH, Razafimanjato H and Heraud JM (2020). Genetic diversity and molecular epidemiology of respiratory syncytial virus circulated in Antananarivo, Madagascar, from 2011 to 2017: Predominance of ON1 and BA9 genotypes. J Clin Virol 129: 104506.

57. Reis J and Shaman J (2016). Retrospective Parameter Estimation and Forecast of Respiratory Syncytial Virus in the United States. PLoS Comput Biol 12(10): e1005133.

58. Reis J and Shaman J (2018). Simulation of four respiratory viruses and inference of epidemiological parameters. Infectious Disease Modelling 3: 23–34.

59. Rm A (1991). Infectious diseases of humans. Aust J Public Health 16: 208–212.

60. Sapin G, Michault A and Simac C (2001). Seasonal trends of respiratory syncytial virus infections on Reunion Island gathering data among hospitalized children. Bulletin de la Societe de Pathologie Exotique (1990) 94(1): 3-4.

61. Shi T, McAllister DA, O’Brien KL, Simoes EA, Madhi SA, Gessner BD, Polack FP, Balsells E, Acacio S and Aguayo C (2017). Global, regional, and national disease burden estimates of acute lower respiratory infections due to respiratory syncytial virus in young children in 2015: a systematic review and modelling study. The Lancet 390(10098): 946–958.

62. Wambua J, Munywoki PK, Coletti P, Nyawanda BO, Murunga N, Nokes DJ and Hens N (2022). Drivers of respiratory syncytial virus seasonal epidemics in children under 5 years in Kilifi, coastal Kenya. PLoS One 17(11): e0278066.

63. Weber A, Weber M and Milligan P (2001). Modeling epidemics caused by respiratory syncytial virus (RSV). Math Biosci 172(2): 95–113.

64. Wesolowski A, Mensah K, Brook CE, Andrianjafimasy M, Winter A, Buckee CO, Razafindratsimandresy R, Tatem AJ, Heraud J-M and Metcalf CJE (2016). Introduction of rubella-containing-vaccine to Madagascar: implications for roll-out and local elimination. Journal of the Royal Society Interface 13(117): 20151101.

65. White L, Mandl J, Gomes M, Bodley-Tickell A, Cane P, Perez-Brena P, Aguilar J, Siqueira M, Portes S and Straliotto S (2007). Understanding the transmission dynamics of respiratory syncytial virus using multiple time series and nested models. Math Biosci 209(1): 222–239.

66. WHO (2002). WHO Manual on Animal Influenza Diagnosis and Surveillance. 105.

67. WHO (2011). Manual for the laboratory diagnosis and virological surveillance of influenza, World Health Organization.

68. WHO (2013). Global epidemiological surveillance standards for influenza. 84.

69. Wood SN (2001). mgcv: GAMs and generalized ridge regression for R. R news 1(2): 20–25.

