## Supplementary Appendix for "Climatic drivers of seasonal dynamics for Respiratory Syncytial Virus (RSV) in Antananarivo, Madagascar, 2011-2021"

**Text S1. Laboratory Details.**

Nasopharyngeal swabs were screened for influenza A and B, RSV and, since March 2020, SARS-CoV-2 using discrete real-time RT-PCR assays. This file reports the protocol for the detection of RSV by real-time RT-PCR.

*Text S1.1 Total Nucleic Acid Extraction*

For the study, viral RNA and DNA were extracted from 150 μL aliquots of each sample, using the Nucleospin® Dx Virus (MACHEREY-NAGEL) kit according to manufacturer's protocol. Whenever possible, samples were subject to RT-PCR testing immediately following extraction. In exceptional cases, extracted RNA and DNA were stored at –80°C following extraction and prior to subsequent molecular analysis.

*Text S1.2 RSV RT-PCR assay*

The RSV RT-PCR assay primers and probe were designed with the aid of Primer Express (Applied Biosystems) and Beacon Designer (Premier Biosoft International) to conserved regions of the matrix gene identified from alignments of sequences available in GenBank (M74568, M11486, AY911262, U39662, NC_001781, AF013254, AY353550).

The primer and probe sequences used were as follows:
forward primer 5'-GGCAAATATGGAAACATACGTGAA-3';
reverse primer 5'-TCTTTTTCTAGGACATTGTAYTGAACAG-3'; and
probe 5'-CTGTGTATGTGGAGCCTTCGTGAAGCT-3'.

The probe was labeled at the 5'end with FAM and quenched at the 3'end with Black Hole Quencher-1 (Biosearch Technologies) (Fry *et al.* 2010).

For the detection of RSV, we performed RT-PCR as a single step using the Superscript III Platinum One-step Quantitative RT-PCR System (Invitrogen, California, USA). Briefly, the reaction mixture contained 12.5μL of 2x Reaction Mix, 0.2μM of each primer and probe, 2 mM of MgS04, 0.6µL of Superscript III RT Platinum TaqMix and nuclease-free water for a final volume of 22.5µL. Then, 2.5µL of RNA extract were added to each tube.

The thermal cycling was implemented on a QuantStudio™ 5 Real-Time PCR System (Applied Biosystem) with a reverse transcription step at $50^0$C for 30 min followed by a 2 min hold at $95^0$C. Amplification consisted in 45 cycles of denaturation at 95°C for 15 seconds and annealing-extension at 55°C for 30 sec. Each specimen was also tested for the human ribonuclease P gene to ensure RNA extract integrity.

**Supplementary Figures:**

**Figure S1. Time series of reported febrile cases and corresponding RSV infection in Antananarivo (2011-2021).** Lines are colored by reporting hospital according to legend, with CENHOSOA reporting most consistently across the time series. Solid lines correspond to the number of febrile samples received and dashed lines to the corresponding subset testing positive for RSV infection.

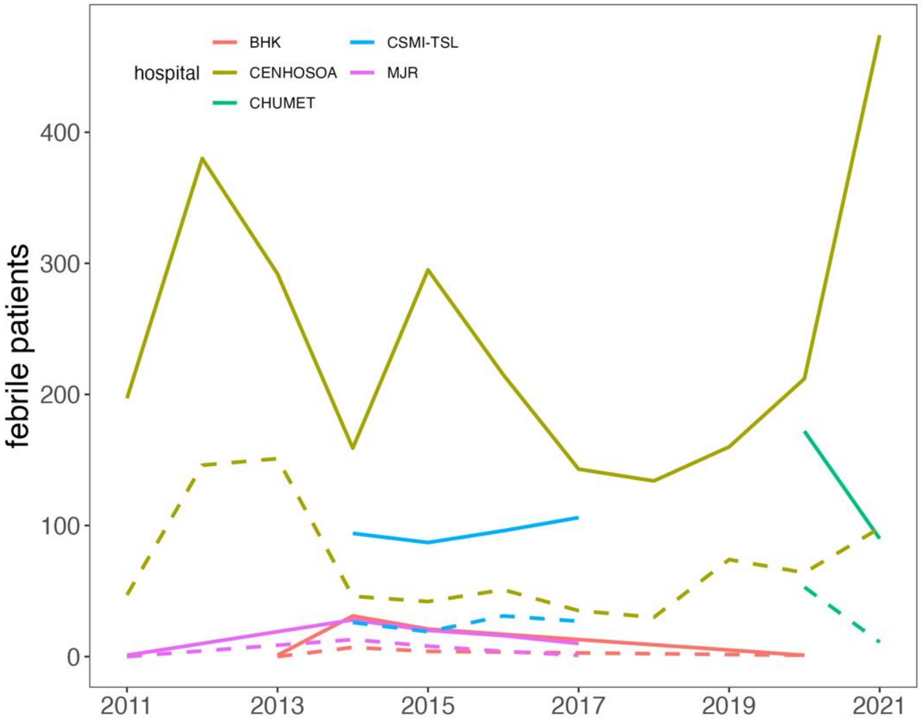

**Figure S2. Outputs from TSIR fitting to weekly RSV cases in Antananarivo, 2011-2021.** (**A**) Susceptible reconstruction from cumulative reported weekly RSV cases (X, red), against cumulative weekly births (Y, green) for the catchment area across the study period and predictions (Yhat, blue) from linear regression. (**B**) Corresponding reporting rate ($\rho$) estimated from the slope of regression in panel **A**, and (**C**) reconstructed deviations (Z) around the (**D**) mean estimated susceptible population through time. (**E**) Fitted weekly transmission rate for RSV by week of year (values held constant at 2-week intervals to avoid overfitting), with shading corresponding to 95% confidence intervals by standard error. Bottom panels show the data (blue) against (**F**) 10 randomly chosen stochastic simulations (red) and (**G**) the (inverse) data against mean of the simulations with confidence intervals. All plots were generated in the R package tsiR (Becker *et al.* 2017). Metrics from TSIR fitting are reported in Table S3.

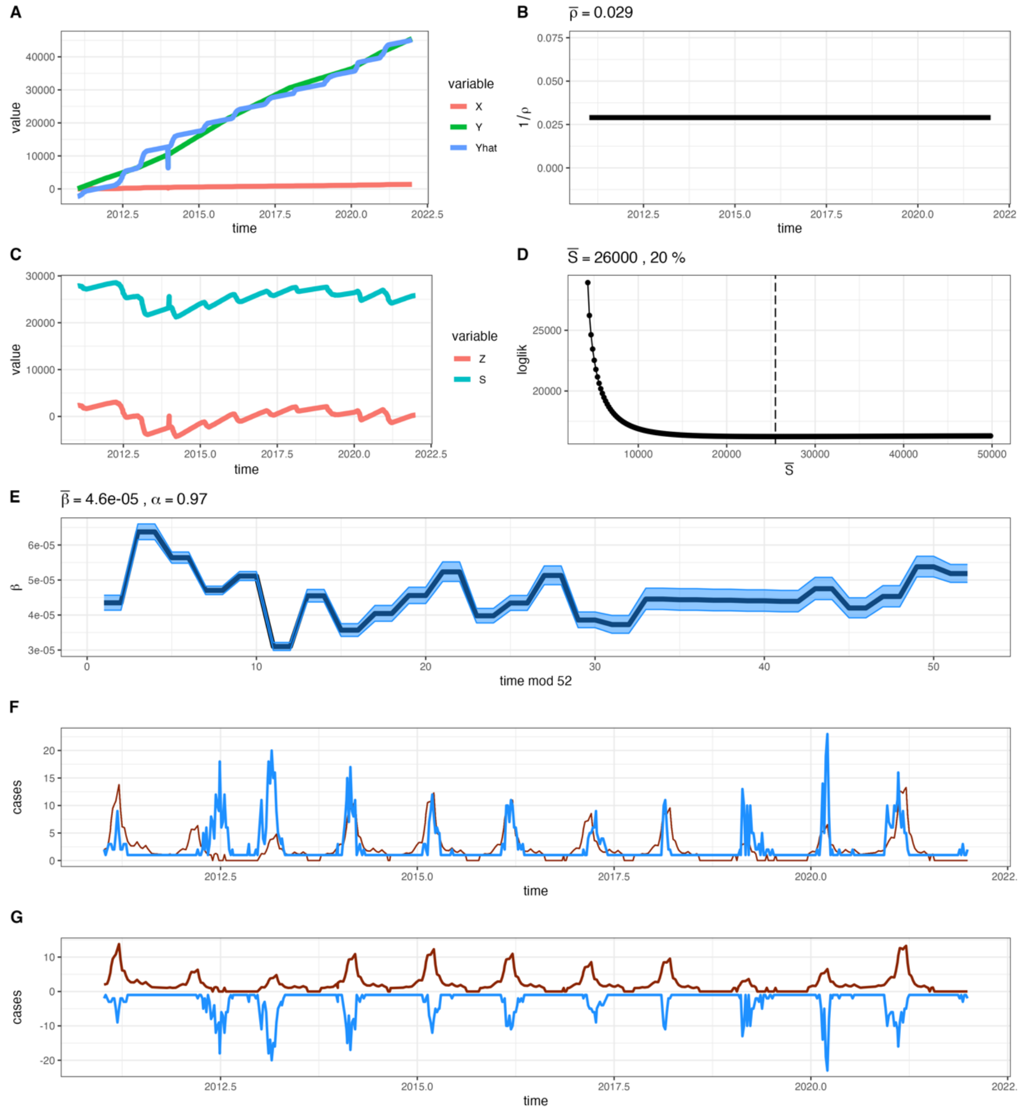

**Figure S3: Inter- and intra-annual trends in climate and RSV case counts from GAMs.** All panels depict inter-annual numerical linear trend (left column), intra-annual numerical trend (middle column), and annual deviations from overall trend (right column) for weekly (**A**) total precipitation (mm), (**B**) relative humidity (%), (**C**) mean temperature ($^{0}$C), and (**D**) RSV case counts. The left and middle columns show outputs derived from GAMs 1A-4A in Table S5 while the right column captures outputs derived from GAMs 1B-4B in Table S5. For left and middle columns, mean estimates are shown as solid lines with gray shading corresponding to 95% confidence intervals by standard error. For right columns, boxplots show mean output and 95% confidence intervals (CI) by standard error. An effect is shaded in gray if the 95% CI crosses zero across the entire range of the predictor variable; in contrast, an effect is shaded in blue and considered "significant" if the 95% CI does not cross zero. GAMs were computed in the gaussian family for panels **A-C** and in the poisson family for panel **D**.

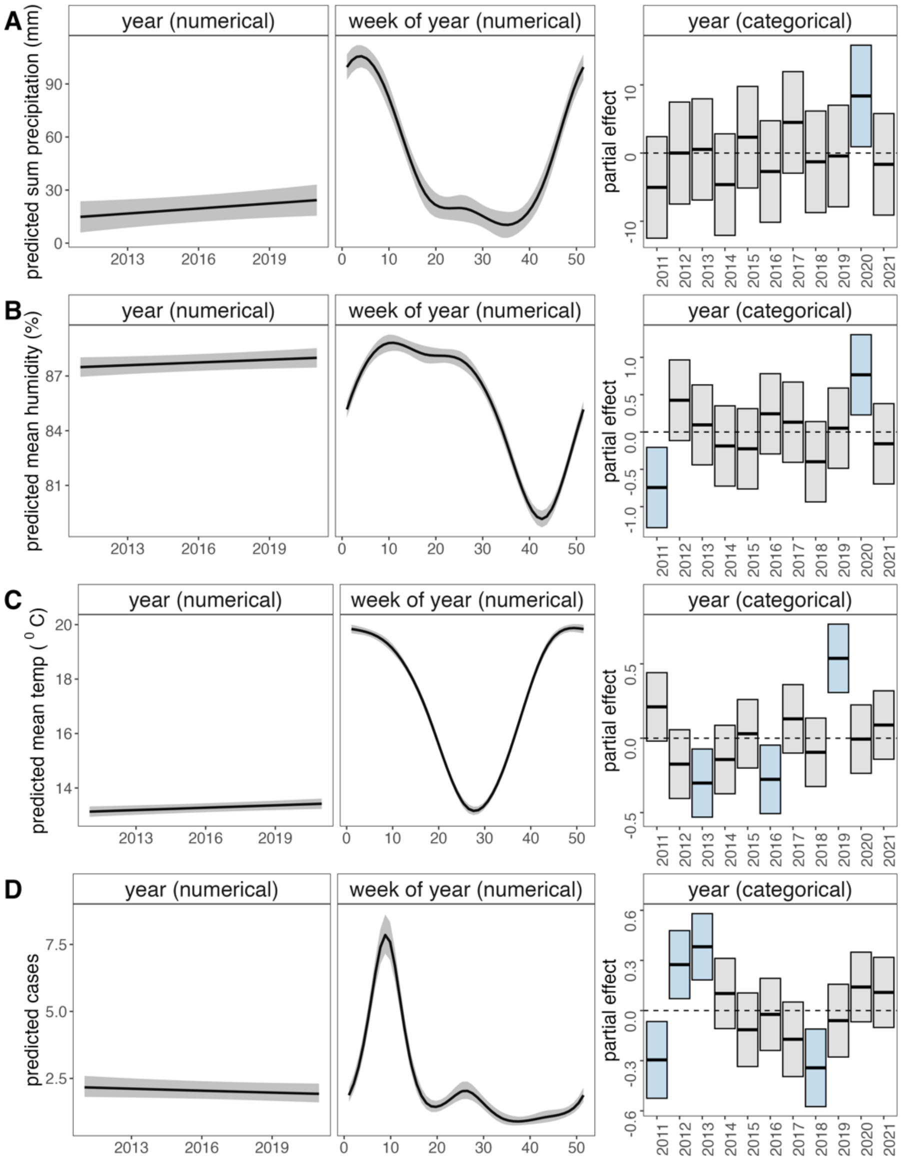

**Supplementary Tables:**

**Table S1: GAM output from correlates of RSV infection analysis**

| **GAM 1** | | | |
|---|---|---|---|
| **Formula:** | RSV ~ s(doy, bs = "cc") + s(year, bs = "re") + s(age, bs = "tp") + s(hospital, bs = "re") + s(sex, bs = "re") | | |
| **$R^2$:** 0.284; **Deviance Explained:** 26.7%; **AIC:** 2886.86 | | | |
| **smooth term** | **edf** | **chi-sq.** | **p-value** |
| day of year | 6.22 | 2205.83 | <0.001*** |
| year | 9.03 | 147.70 | <0.001*** |
| age | 6.01 | 177.25 | <0.001*** |
| hospital | 2.21 | 56.09 | <0.001*** |
| sex | 0.539 | 1.16 | 0.146 |

| **GAM 2** | | | |
|---|---|---|---|
| **Formula** | RSV ~ s(doy, bs = "cc") + s(year, bs = "tp") + s(age, bs = "tp") + s(hospital, bs = "re") + s(sex, bs = "re") | | |
| **$R^2$:** 0.281; **Deviance Explained:** 26.6%; **AIC:** 2890.91 | | | |
| **smooth term** | **edf** | **chi-sq.** | **p-value** |
| day of year | 6.16 | 388.15 | <0.001*** |
| year | 8.47 | 133.22 | <0.001*** |
| age | 5.98 | 175.84 | <0.001*** |
| hospital | 2.12 | 9.69 | 0.002** |
| sex | 0.50 | 1.00 | 0.157 |

| **GAM 3** | | | |
|---|---|---|---|
| **Formula** | RSV ~ s(doy, bs = "cc") + s(year, bs = "re") | | |
| **$R^2$:** 0.205; **Deviance Explained:** 19.8%; **AIC:** 3339.23 | | | |
| **smooth term** | **edf** | **chi-sq.** | **p-value** |
| day of year | 6.31 | 2758.3 | <0.001*** |

| | | | |
|---|---|---|---|
| year | 9.36 | 188.2 | <0.001*** |

significance at p-value <0.001***, <0.01**, <0.1*

**Table S2. Model comparison for FOI fitting**

| Number of Parameters Estimated | Model FOI Structure | Negative log-likelihood | AIC | Model converged? |
|---|---|---|---|---|
| 1 | One constant FOI | 5075 | 10152.01 | yes |
| 2 | 0-5; 5+ | 392.55 | 789.1 | yes |
| | 0-4; 4+ | 289.19 | 582.38 | yes |
| | 0-3; 3+ | 238.84 | 481.68 | yes |
| | 0-2; 2+ | 285.67 | 575.33 | yes |
| | 0-1; 1+ | 612.47 | 1228.93 | yes |
| | 0-.5; .5+ | 10-59.39 | 2122.78 | yes |
| 3 | 0-1;1-2;2+ | 269.71 | 545.41 | no |
| 4 | 0-1;1-2;2-3;3+ | 230.41 | 468.81 | yes |
| 5 | 0-1;1-2;2-3;3-4;4+ | 220.37 | 450.73 | yes |
| 6 | 0-1;1-2;2-3;3-4;4-5;5+ | 220.15 | 452.3 | yes |
| | 0-.5;.5-1;1-2;2-3;3-4;4+ | 215.45 | 442.91 | yes |
| 14 | 0-.5;.5-1;1-2;2-3;3-4;4-10;10-20;20-30;30-40;40-50;50-60;60-70;70-80; 80+ | 171.02 | 370.04 | no |
| 13 | 0-.5;.5-1;1-2;2-3;3-4;4-10;10-20;20-30;30-40;40-50;50-60;60-70+ | 171.26 | 368.51 | yes |
| 12 | 0-.5;.5-1;1-2;2-3;3-4;4-10;10-20;20-30;30-40;40-50;50-60;60+ | 174.79 | 373.58 | yes |
| 12 | 0-.5;.5-1;1-2;2-3;3-4;4-10;10-30;30-40;40-50;50-60;60-70;70+ | 171.27 | 366.54 | yes |
| 11 | 0-.5;.5-1;1-2;2-3;3-4;4-10;10-30;30-50;50-60;60-70;70+ | 171.28 | 364.56 | yes |
| 10 | 0-.5;.5-1;1-2;2-3;3-4;4-10;10-30;30-60;60-70;70+ | 172.29 | 364.58 | yes |
| 9 | 0-.5;.5-1;1-2;2-3;3-4;4-10;10-30;30-70;70+ | 176.76 | 371.51 | yes |
| 9 | 0-.5;.5-1;1-2;2-3;3-4;4-10;10-60;60-70;70+ | 173.48 | 364.96 | yes |
| 8 | 0-.5;.5-1;1-2;2-3;3-4;4-60;60-70;70+ | 176.04 | 368.07 | yes |
| 9 | 0-.5;.5-1;1-2;2-3;3-4;4-5;5-60;60-70;70+ | 174.8 | 367.6 | yes |
| 9 | 0-.5;.5-1;1-2;2-3;3-4;4-20;20-60;60-70;70+ | 172.29 | 362.58 | yes |
| 10 | 0-.5;.5-1;1-2;2-3;3-4;4-10;10-20;20-60;60-70;70+ | 172.26 | 364.51 | yes |

| | | | | |
|---|---|---|---|---|
| 9 | 0-.5;.5-1;1-2;2-3;3-4;4-10;10-15;15-60;60-70;70+ | 172.7 | 363.39 | yes |
| **9** | **0-.5;.5-1;1-2;2-3;3-4;4-25;25-60;60-70;70+** | **172.19** | **362.38** | **yes** |
| 9 | 0-.5;.5-1;1-2;2-3;3-4;4-30;30-60;60-70;70+ | 172.81 | 363.61 | yes |
| 91 | Unique FOI for all age classes | 167.22 | 516.45 | no |

**FOI Estimates from Best Fit Model (*highlighted above in red*)**

| **Lower Age Bin*** | **Upper Age Bin*** | **FOI** | **FOI (95% lower CI)** | **FOI (95% upper CI)** |
|---|---|---|---|---|
| 0 | 0.5 | 1.03 | 0.92 | 1.13 |
| 0.5 | 1 | 1.5 | 1.27 | 1.73 |
| 1 | 2 | 0.75 | 0.56 | 0.95 |
| 2 | 3 | 0.48 | 0.21 | 0.75 |
| 3 | 4 | 0.63 | 0.37 | 0.88 |
| 4 | 25 | 0.04 | 0.03 | 0.05 |
| 25 | 60 | 0.02 | 0.01 | 0.03 |
| 60 | 70 | 0.11 | 0.05 | 0.16 |
| 70 | 89 | 0.33 | 0.15 | 0.51 |

**Age bins are bound to values equal to or greater than the lower bounds and less than the upper bounds listed here.*

**Table S3: TSIR model output to estimate weekly RSV transmission rate**

| **A. Regression for Susceptible Reconstruction** | | | | |
|---|---|---|---|---|
| **Formula:** cumulative_weekly_births ~ cumulative_weekly_RSV_cases | | | | |
| **$R^2$:** 0.983; **F-Statistic:** 32830; **p-value:** <0.001 | | | | |
| **Term** | **Estimate** | **95% CI** | **T-value** | **P-value** |
| Intercept | -2344.9 | [-2652.3- -2037.6] | -14.95 | <0.001*** |
| cumulative_weekly_ RSV_cases | 34.5 | [34.2-34.9] | 181.2 | <0.001*** |
| significance at p-value <0.001***, <0.01**, <0.1* | | | | |

| **B. Log-Link Regression for Transmission Estimation** |
|---|
| **Formula:** Inew ~ -1 + as.factor(period) + offset(alpha*lIminus) + offset(lSminus), family="poisson" |
| **Pseudo-$R^2$ McFadden:** 0.641; |

| **C. Weekly Transmission Rate Estimates** | | | | |
|---|---|---|---|---|
| **Biweek** | **Weeks** | $\beta$ | $\beta$ **(lower 95% CI)** | $\beta$ **(upper 95% CI)** |
| 1 | 1-2 | $4.35x10^{-5}$ | $4.14\ x10^{-5}$ | $4.57\ x10^{-5}$ |
| 2 | 3-4 | $6.37\ x10^{-5}$ | $6.15\ x10^{-5}$ | $6.60\ x10^{-5}$ |
| 3 | 5-6 | $5.64\ x10^{-5}$ | $5.48\ x10^{-5}$ | $5.80\ x10^{-5}$ |
| 4 | 7-8 | $4.70\ x10^{-5}$ | $4.58\ x10^{-5}$ | $4.83\ x10^{-5}$ |
| 5 | 9-10 | $5.11\ x10^{-5}$ | $4.99\ x10^{-5}$ | $5.24\ x10^{-5}$ |
| 6 | 11-12 | $3.10\ x10^{-5}$ | $2.99\ x10^{-5}$ | $3.22\ x10^{-5}$ |
| 7 | 13-14 | $4.55\ x10^{-5}$ | $4.37\ x10^{-5}$ | $4.73\ x10^{-5}$ |
| 8 | 15-16 | $3.57\ x10^{-5}$ | $3.39\ x10^{-5}$ | $3.75\ x10^{-5}$ |
| 9 | 17-18 | $4.04\ x10^{-5}$ | $3.82\ x10^{-5}$ | $4.27\ x10^{-5}$ |
| 10 | 19-20 | $4.56\ x10^{-5}$ | $4.33\ x10^{-5}$ | $4.79\ x10^{-5}$ |
| 11 | 21-22 | $5.23\ x10^{-5}$ | $4.96\ x10^{-5}$ | $5.52\ x10^{-5}$ |
| 12 | 23-24 | $3.98\ x10^{-5}$ | $3.77\ x10^{-5}$ | $4.19\ x10^{-5}$ |
| 13 | 25-26 | $4.34\ x10^{-5}$ | $4.13\ x10^{-5}$ | $4.56\ x10^{-5}$ |
| 14 | 27-28 | $5.13\ x10^{-5}$ | $4.87\ x10^{-5}$ | $5.40\ x10^{-5}$ |
| 15 | 29-30 | $3.86\ x10^{-5}$ | $3.64\ x10^{-5}$ | $4.09\ x10^{-5}$ |
| 16 | 31-32 | $3.73\ x10^{-5}$ | $3.48\ x10^{-5}$ | $3.99\ x10^{-5}$ |

| | | | | |
|---|---|---|---|---|
| 17 | 33-34 | 4.46 x$10^{-5}$ | 4.16 x$10^{-5}$ | 4.77 x$10^{-5}$ |
| 18 | 35-36 | 4.44 x$10^{-5}$ | 4.14 x$10^{-5}$ | 4.75 x$10^{-5}$ |
| 19 | 37-38 | 4.42 x$10^{-5}$ | 4.12 x$10^{-5}$ | 4.74 x$10^{-5}$ |
| 20 | 39-40 | 4.41 x$10^{-5}$ | 4.11 x$10^{-5}$ | 4.72 x$10^{-5}$ |
| 21 | 41-42 | 4.39 x$10^{-5}$ | 4.09 x$10^{-5}$ | 4.70 x$10^{-5}$ |
| 22 | 43-44 | 4.75 x$10^{-5}$ | 4.44 x$10^{-5}$ | 5.08 x$10^{-5}$ |
| 23 | 45-46 | 4.20 x$10^{-5}$ | 3.92 x$10^{-5}$ | 4.49 x$10^{-5}$ |
| 24 | 47-48 | 4.53 x$10^{-5}$ | 4.23 x$10^{-5}$ | 4.84 x$10^{-5}$ |
| 25 | 49-50 | 5.38 x$10^{-5}$ | 5.09 x$10^{-5}$ | 5.68 x$10^{-5}$ |
| 26 | 51-52 | 5.18 x$10^{-5}$ | 4.93 x$10^{-5}$ | 5.44 x$10^{-5}$ |

**Table S4: GLM output from climate correlates of RSV transmission**

**A. Climate model comparisons**

| | Estimate | | | | | | | |
|---|---|---|---|---|---|---|---|---|
| **Model** | **Intercept** | **Mean Humidity** | **Mean Temp** | **Sum Precip.** | **DF** | **Log-Lik** | **AICc** | **δAICc** |
| **1** | **-9.79** | **-0.0041** | **0.0058** | **0.00074** | **5** | **309.16** | **-608.22** | **0** |
| 2 | -9.56 | -0.0058 | NA | 0.00094 | 4 | 307.45 | -606.83 | 1.39 |
| 3 | -10.2 | NA | 0.0093 | 0.00055 | 4 | 306.61 | -605.14 | 3.08 |
| 4 | -10.1 | NA | NA | 0.0008 | 3 | 300.9 | -595.77 | 12.46 |
| 5 | -10.3 | NA | 0.015 | NA | 3 | 299.17 | -592.3 | 15.9 |
| 6 | -10.3 | 8.40*10-5 | 0.015 | NA | 4 | 299.17 | -590.27 | 17.9 |
| 7 | -9.81 | -0.0023 | NA | NA | 3 | 280.68 | -555.32 | 52.9 |
| 8 | -10 | NA | NA | NA | 2 | 279.64 | -555.26 | 53 |

**B. Best fit model summary (#1)**

**Formula:** log_beta ~ mean_humid + sum_precip + mean_temp

**$R^2$:** 0.098; **F-Statistic:** 20.59; **p-value:** <0.001

| **Term** | **Estimate** | **95% CI** | **T-value** | **P-value** |
|---|---|---|---|---|
| Intercept | -9.79 | [-10.2- -9.43] | -53.2 | <0.001*** |
| Mean Humidity | -0.0041 | [-0.0077-0.00055] | -2.26 | 0.024* |
| Mean Temp | 0.0058 | [-0.00035-0.12] | 4.49 | <0.001*** |
| Sum Precip. | 0.00074 | [0.00042-0.0011] | 1.85 | 0.065* |

significance at p-value <0.001***, <0.01**, <0.1*

**Table S5: GAM output from interannual climate trend analysis**

| | | | |
|---|---|---|---|
| **GAM 1A: Precipitation** | | | |
| **Formula:** | sum_weekly_precip ~ year + s(week, k = 7, bs = "cc") | | |
| **$R^2$:** 0.483; **Deviance Explained:** 48.8% | | | |
| **parametric coefficient** | **estimate [95% CI]** | **t-value** | **p-value** |
| Year | 0.945 [0.022-1.87] | 2.01 | <0.001*** |
| **smooth term** | **edf** | **F-stat** | **p-value** |
| week | 4.56 | 10-5.8 | <0.001*** |
| **GAM 1B: Precipitation** | | | |
| **Formula:** | sum_weekly_precip ~ s(week, k = 7, bs = "cc") + s(year, k = 7, bs = "re") | | |
| **$R^2$:** 0.491 **Deviance Explained:** 50% | | | |
| **smooth term** | **edf** | **F-stat** | **p-value** |
| week | 4.56 | 107.5 | <0.001*** |
| year | 5.43 | 1.19 | 0.0172* |
| **GAM 2A: Humidity** | | | |
| **Formula** | mean_weekly_humidity ~ year + s(week, k = 7, bs = "cc") | | |
| **$R^2$:** 0.708; **Deviance Explained:** 71.1% | | | |
| **parametric coefficient** | **estimate [95% CI]** | **t-value** | **p-value** |
| year | 0.051 [-0.004-0.10-5] | 1.83 | 0.068* |
| **smooth term** | **edf** | **F-stat** | **p-value** |
| week | 4.84 | 275.7 | <0.001*** |
| **GAM 2B: Humidity** | | | |
| **Formula:** | mean_weekly_humidity ~ s(week, k = 7, bs = "cc") + s(year, k = 7, bs = "re") | | |
| **$R^2$:** 0.72 **Deviance Explained:** 72.6% | | | |
| **smooth term** | **edf** | **F-stat** | **p-value** |
| week | 4.85 | 288.2 | <0.001*** |
| year | 7.37 | 2.80 | <0.001*** |

| **GAM 3A: Temperature** | | | |
|---|---|---|---|
| **Formula** | mean_weekly_temperature ~ year + s(week, k = 7, bs = "cc") | | |
| **$R^2$:** 0.914; **Deviance Explained:** 91.5% | | | |
| **parametric coefficient** | **estimate [95% CI]** | **t-value** | **p-value** |
| year | 0.029 [0.010-0.048] | 2.93 | 0.0035** |
| **smooth term** | **edf** | **F-stat** | **p-value** |
| week | 4.86 | 1207 | <0.001*** |

| **GAM 3B: Temperature** | | | |
|---|---|---|---|
| **Formula:** | mean_weekly_temperature ~ s(week, k = 7, bs = "cc") + s(year, k = 7, bs = "re") | | |
| **$R^2$:** 0.922 **Deviance Explained:** 92.4% | | | |
| **smooth term** | **edf** | **F-stat** | **p-value** |
| week | 4.87 | 1338.3 | <0.001*** |
| year | 8.74 | 6.93 | <0.001*** |

| **GAM 4A: RSV Cases** | | | |
|---|---|---|---|
| **Formula** | total_weekly_cases ~ year + s(week, k = 7, bs = "cc") | | |
| **$R^2$:** 0.352; **Deviance Explained:** 46.8% | | | |
| **parametric coefficient** | **estimate [95% CI]** | **z-value** | **p-value** |
| year | -0.012 [-0.029-0.0049] | -1.39 | 0.165 |
| **smooth term** | **edf** | **chi-sq.** | **p-value** |
| week | 4.95 | 720.3 | <0.001*** |

| **GAM 4B: RSV Cases** | | | |
|---|---|---|---|
| **Formula:** | total_weekly_cases ~ s(week, k = 7, bs = "cc") + s(year, k = 7, bs = "re") | | |
| **$R^2$:** 0.403 **Deviance Explained:** 52.1% | | | |
| **smooth term** | **edf** | **chi-sq.** | **p-value** |
| week | 4.95 | 720.9 | <0.001*** |
| year | 8.78 | 72.8 | <0.001*** |

significance at p-value <0.001***, <0.01**, <0.1*

### Supplementary References


1. Becker AD and Grenfell BT (2017). tsiR: An R package for time-series Susceptible-Infected-Recovered models of epidemics. PLoS One 12(9): e0185528.

2. Fry AM, Chittaganpitch M, Baggett HC, Peret TC, Dare RK, Sawatwong P, Thamthitiwat S, Areerat P, Sanasuttipun W and Fischer J (2010). The burden of hospitalized lower respiratory tract infection due to respiratory syncytial virus in rural Thailand. PLoS One 5(11): e15098.
